# Transcriptomics-based subphenotyping of the human placenta enabled by weighted correlation network analysis in early-onset preeclampsia with and without fetal growth restriction

**DOI:** 10.1101/2023.03.08.23286998

**Authors:** William E. Ackerman, Catalin S. Buhimschi, Thomas L. Brown, Guomao Zhao, Taryn L. Summerfield, Irina A. Buhimschi

## Abstract

**Background:** Placental disorders contribute to pregnancy complications, including preeclampsia (PE) and fetal growth restriction (FGR), but debate regarding their specific pathobiology persists. Our objective was to apply transcriptomics with weighted gene correlation network analysis (WGCNA) to further clarify the placental dysfunction in these conditions.

**Methods:** We performed RNA sequencing with WGNCA using human placental samples (n=30), separated into villous tissue and decidua basalis, and clinically grouped as follows: (1) early-onset PE (EOPE)+FGR (n=7); (2) normotensive, nonanomalous preterm FGR (n=5); (2) EOPE without FGR (n=8); (4) spontaneous idiopathic preterm birth (PTB, n=5) matched for gestational age (GA); and (5) uncomplicated term births (TB, n=5). Our data was compared with RNA-seq datasets from public databases (GSE114691, GSE148241, and PRJEB30656; n=130 samples).

**Results:** We identified 14 correlated gene modules in our specimens, of which most were significantly correlated with birthweight and maternal blood pressure. Of the 3 network modules consistently predictive of EOPE±FGR across datasets, we prioritized a co-expression gene group enriched for hypoxia-response and metabolic pathways for further investigation. Cluster analysis based on transcripts from this module and the glycolysis/gluconeogenesis metabolic pathway consistently distinguished a subset of EOPE±FGR samples with an expression signature suggesting modified tissue bioenergetics. We demonstrated that the expression ratios of *LDHA*/*LDHB* and *PDK1*/*GOT1* could be used as surrogate indices for the larger panels of genes in identifying this subgroup.

**Conclusions:** We provide novel evidence for a molecular subphenotype consistent with a glycolytic metabolic shift that occurs more frequently but not universally in placental specimens of EOPE±FGR.

**Graphical Abstract:** 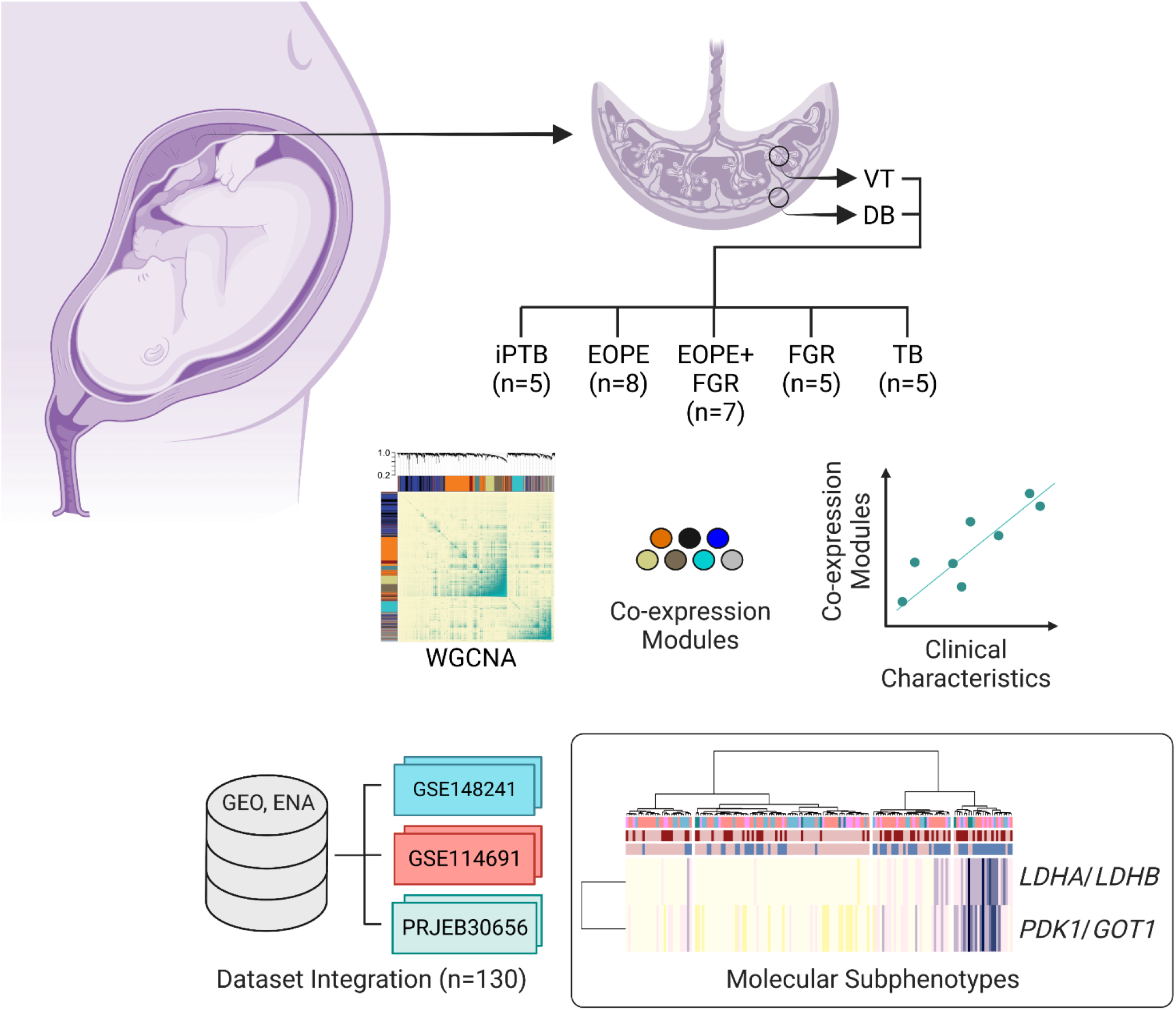

## Introduction

Placental function within physiological limits is essential for successful pregnancy outcomes. Abnormal development or dysfunction of this vital organ can lead to severe pregnancy complications, inclusively referred to as pregnancy-related “placental syndromes” ^1, 2^. Disorders associated with disordered placentation include fetal growth restriction (FGR) and preeclampsia (PE) ^2^, both of which increase the acute risk of perinatal morbidity and mortality ^1, 3, 4^. These disorders can predispose pregnant individuals and their offspring to chronic health problems ^1^.

PE has traditionally been defined clinically by new-onset or worsening hypertension during pregnancy >20 weeks with accompanying proteinuria or other signs of organ system involvement ^5^. FGR, which can accompany PE or occur independently, is typically detected prior to birth through ultrasonographic estimation of fetal weight. When it occurs, placental insufficiency is strongly implicated in the pathophysiology of FGR and PE alike; however, the precise etiological underpinnings of these complex conditions remain incompletely understood ^2, 3, 6^. “Final common pathway” models for both disease states have been proposed. These posit that diverse etiologies can converge to produce suboptimal uteroplacental perfusion, which in turn may manifest itself in stereotyped, albeit inconsistent, clinical findings ^6^.

Weighted gene correlation network analysis (WGCNA) is a data reduction technique applied widely in gene expression studies to identify and functionally categorize clusters of highly correlated transcripts (co-expression modules) ^7^. The resulting clusters can then be related to clinical variables (e.g., blood pressure) using consolidation metrics such as “eigengenes” ^8^, and “metagenes” ^9^. Since modules thus constructed are impartial to clinical outcome, WGCNA can be applied when analyzing datasets with unknown sample stratifications. Translational examples include use in identifying tumor subgroups with differing outcomes but common presentations ^10^. By distinguishing molecular subtypes and their links to candidate prognostic markers and druggable targets, this approach allows investigators to crystalize transcriptome expression data in a potentially actionable way ^7^.

Recognizing the considerable heterogeneity in outcomes for patients presenting with PE and FGR, Cox and colleagues utilized unsupervised clustering approaches for class discovery using gene expression profiling in a large cohort of placental samples from affected pregnancies ^11, 12^. Their data-driven classifications revealed several robust subclusters that differed in clinical and histopathologic findings as well as molecular-level functional enrichments. These studies illustrated the potential for functional genomics in molecular phenotyping as an adjunct to clinical diagnostic criteria in classifying PE and FGR and offered new insights into the pathoetiology of these conditions.

Inspired by these findings, we utilized WGCNA in combination with transcriptome profiling by RNA sequencing (RNA-seq) to: (1) define other possible placental subphenotypes in PE and FGR; and (2) further elucidate dysregulated placental processes associated with these syndromes.

## Methods

The data that support the findings of this study are available from the corresponding author upon reasonable request. The RNA-seq data generated for this publication have been deposited in NCBI’s Gene Expression Omnibus (GSE203507). Detailed Methods are provided in the Supplemental Material.

### Study Approval

This study was approved by the IRBs of Yale University, The Ohio State University, the Abigail Wexner Research Institute at Nationwide Children’s Hospital, and the University of Illinois at Chicago. All study participants provided written informed consent.

### Recruitment and Tissue Collection

Transcriptomics was performed using placentas from 30 pregnant individuals carefully phenotyped and categorized clinically: (1) early-onset PE (EOPE)+FGR (n=7); (2) normotensive, nonanomalous early-onset FGR (n=5); (3) EOPE without FGR (n=8); (4) spontaneous idiopathic preterm birth (PTB, n=5) without FGR or triple I (intrauterine infection and/or inflammation), and matched for gestational age (GA); and (5) uncomplicated term births (TBs, n=5). Data generated from specimens comprising the PTB and TB control groups have been previously analyzed ^13–15^ but were reevaluated starting from the raw data. A summary of these samples is presented in Tables S1&S2.

### RNA Extraction and Bulk RNA Sequencing (RNA-seq)

Villous tissue (VT) and basal plate decidua basalis (DB) were collected, and total RNA was isolated, as previously reported ^13, 16^. RNA-seq was performed using the Illumina HiSeq 2500 system.

### Differential Abundance Analysis

Differential transcript abundance was determined using DESeq2 v1.32.0 . Surrogate variable analysis was performed to control for unrecognized heterogeneity. Statistical models included clinical outcome, assigned fetal sex at delivery, and the first 2 components of the surrogate variable analysis as covariates. To minimize batch effects, the RNA-seq data were generated in 2 rounds of sequencing. The PTB and TB specimens reported previously ^13–15^ were isolated and sequenced concurrently with samples S16, S20, S21, and S22 (Table S2).

### Weighted Gene Correlation Network Analysis (WGCNA)

WGCNA was performed using the top quartile (∼4000) of normalized transcript counts exhibiting the greatest variability in each placental region (VT and DB). Correlated gene modules were identified from co-expression networks using the WGCNA R library.

### Module-Trait Associations

Eigengene-based module-trait associations for continuous clinical variables were computed using the WGCNA R package. For categorical variables, we adopted a metagene-centered approach. Briefly, associations between binary outcomes and metagenes, defined as the arithmetic mean of normalized, variance-stabilizing transformed counts for selected transcripts ^9^, were assessed using ROC analysis. For each module, the metagene comprising the top 10 transcripts most positively correlated with select traits was divided by the metagene composed of the top 10 transcripts most negatively correlated with those same traits, yielding a metagene ratio (MGR).

### Comparison with Previously Published Datasets

To assess the consistency of our present findings with those of prior studies, we identified three placenta-derived RNA-seq datasets for further analysis: (1) GSE114691 ^17^, comprising 79 samples; (2) GSE148241 ^18^, consisting of 41 samples; and (3) PRJEB30656 ^19^, consisting of 10 samples (Table S3).

For joint analyses, variance-stabilizing transformed counts were adjusted for study-dependent batch effects.

### Functional Enrichment Analysis

We performed functional enrichment (hypergeometric testing) on prioritized groups of transcripts using clusterProfiler in R.

### qPCR Cross-Validation

We selected 4 transcripts for qPCR cross-validation using the general methodology described previously ^16^: lactate dehydrogenase A (*LDHA*), lactate dehydrogenase B (*LDHB*), pyruvate dehydrogenase kinase 1 (*PDK1*), and glutamic-oxaloacetic transaminase 1 (*GOT1*). These transcripts were chosen based on their relevance to core placental metabolic pathways ^20–23^. TaqMan gene expression assays (Thermo Fisher Scientific) were used for quantification (see the Major Resources Table). Relative abundance was calculated using the comparative Ct method. The geometric mean of the Ct values for β2-microglobulin (*B2M*) and ribosomal protein L30 (*RPL30*) was used as a reference in each reaction, performed in duplicate. Correlations between the expression levels from RNA-seq and qPCR experiments were examined using Pearson’s product-moment correlation (on log-transformed data).

### Statistics

All statistical analyses were performed using a combination of functions and packages within the R v4.1.0, in addition to Prism v9 (GraphPad Software, La Jolla, CA) and MedCalc v20.027 (MedCalc Software Ltd, Ostend, Belgium). Clinical characteristics among the study groups were evaluated using ANOVA or Kruskal–Wallis testing (for continuous variables) and Chi-squared tests (for categorical variables) and for these comparisons, a *p*-value <0.05 was considered statistically significant. Adjustments for multiple comparisons were performed using the Benjamini-Hochberg procedure with FDR<0.1 considered significant. Additional details are provided in the figure legends and Supplemental Material.

## Results

### Overview of Study Population and Differential Abundance Analysis

We performed RNA-seq profiling of paired VT and DB placental samples grouped by clinical phenotype: (1) EOPE+FGR; (2) normotensive, nonanomalous preterm FGR; (3) EOPE without FGR; (4) spontaneous idiopathic preterm birth (PTB) without FGR, matched for GA; and (5) uncomplicated TB. The 2 control groups (PTB and TB) were chosen to account for the changes in gene expression associated with spontaneous preterm delivery or advancing GA. The demographic, clinical, and pregnancy outcome data associated with these placental samples are summarized in Tables S1&S2.

Similarity analysis with hierarchical clustering revealed that PTB and TB placental samples were the most similar groups overall, with EOPE and EOPE+FGR samples forming a more distantly related group and FGR occupying an intermediate position (Fig. 1). Consistently, by principal component (PC) analysis, we noted that PTB and TB placental samples were more closely aggregated than the EOPE, EOPE+FGR, and FGR samples (Fig. 1 B&D). More detailed pairwise comparisons between the groups recapitulated these general observations (Fig. S1-S3).

**Figure 1.**
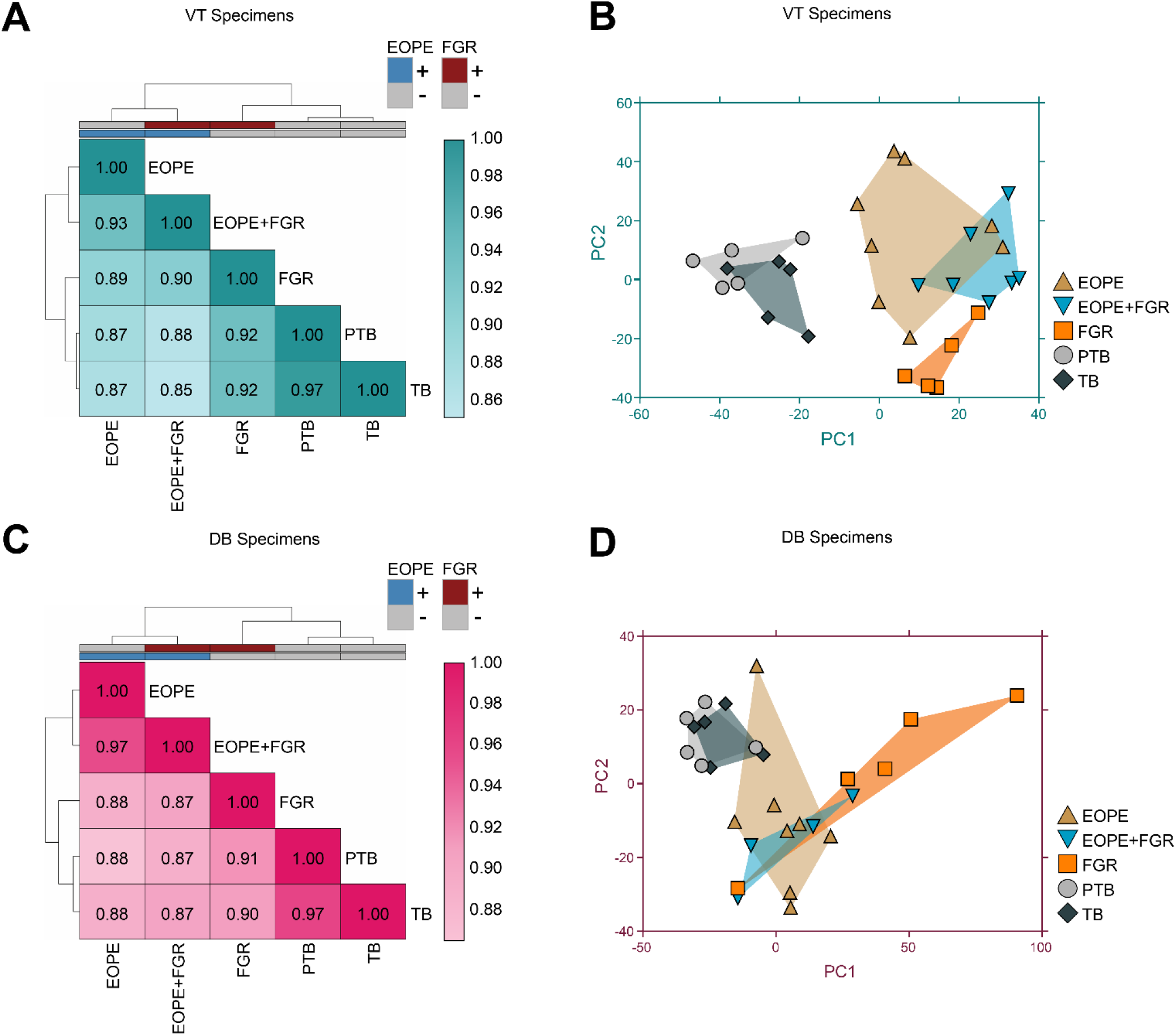
Relationships among samples in clinically-phenotyped groups based on differential transcript abundance analysis. (A) Similarity plot with hierarchical clustering using Ward’s minimum variance method for VT specimens based on the expression of 6,865 consensus differentially abundant transcripts. The color scale is arranged from light (most dissimilar) to dark (identity). (B) Scatterplot showing the results of PC analysis in VT samples. (C) Similarity plot with hierarchical clustering for DB specimens based on the expression of 2,551 consensus differentially abundant transcripts averaged for each clinicallgroup. (D) Scatterplot of PC analysis applied to differentially abundant consensus transcripts in DB specimens. The differentially abundant consensus transcripts for each placental region represent the union of unique transcripts present in at least one pairwise comparison in DESeq2 statistical models that included clinical outcome, assigned fetal sex at delivery, and the first 2 components of the surrogate variable analysis as covariates (FDR<0.1).

### Correlation Network Analysis Reveals Placental Transcript Modules that Correlate with Clinical Variables Semi-independently

We next generated weighted correlation networks to further assess potentially informative gene expression patterns in relation to select clinical variables. This approach enables unsupervised, data-driven assessment of the interplay between molecular signatures and various clinical variables ^7^.

Eight co-expression modules were identified in the VT samples (Fig. 2 A&B, Table S4). Module- trait correlation analysis showed that, among VT samples, modules VT 2 (Blue, 1001 transcripts) and VT 3 (Black, 391 transcripts) were the most strongly correlated with blood pressure, whereas module VT 1 (Orange, 1066 transcripts) showed the strongest correlation with birthweight (Fig. 2C). Modules VT 4 (Turquoise, 327 transcripts) and VT 7 (Brick, 238 transcripts) also exhibited a significant correlation with birthweight in addition to blood pressure, while VT 6 (Gold, 269 transcripts) was correlated with birthweight only (Fig. 2C).

**Figure 2.**
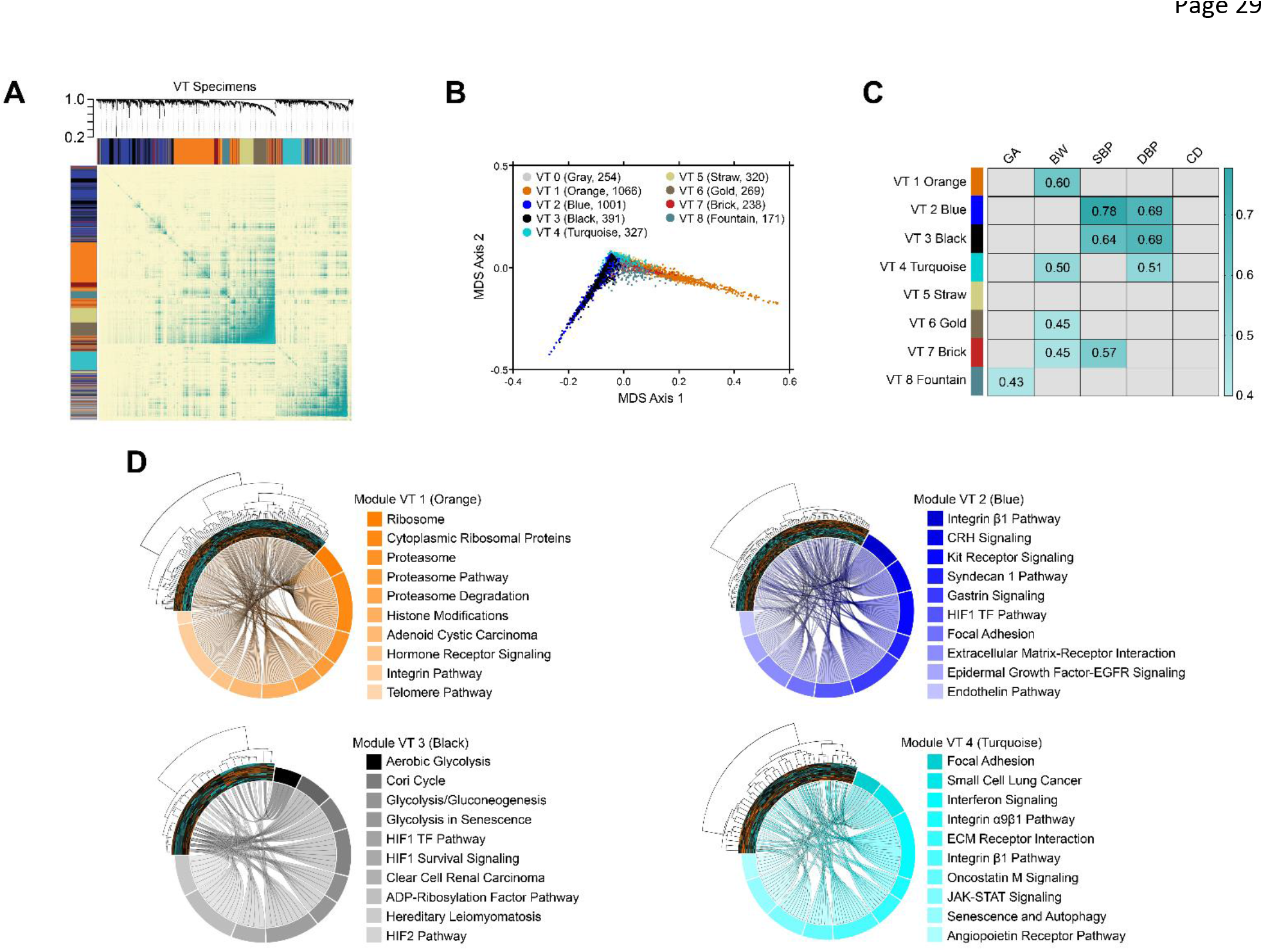
Consensus correlation network modules among villous tissue (VT) placental specimens. WGCNA was applied to the most variable VT transcripts (top quartile, 4,037 RNAs). (A) Clustering dendrograms and topological overlap matrix heatmap (β=9, scale-free topology model fit r^2^=0.90, median connectivity=6.4). Dendrogram colors represent individual transcript modules. (B) Multidimensional scaling was applied to the transcript dissimilarity matrix based on the topological overlap and represented as a scatterplot, color-coded by module. (C) Module-trait correlation analysis. Statistically significant (*p*<0.05) Pearson correlation coefficients are shown in colored boxes. (D) Chord diagrams showing the top10 overrepresented pathways associated with select modules

Functional enrichment analysis of the VT Orange module demonstrated overrepresentation for proteasome pathway mRNAs (including transcripts encoding multiple proteasome subunits) and ribosomal protein subunit transcripts (all FDR<0.006, hypergeometric test) (Fig. 2D). The VT Blue module included numerous PE-associated transcripts curated in the Comparative Toxicogenomics Database, such as *ENG*, *FLT1*, and *LEP* (Table S4), and was enriched (FDR<0.006) for several membrane receptor signaling pathways (Fig. 2D). The VT Black module was chiefly characterized (FDR<0.081) by transcripts associated with the hypoxia-inducible factor (HIF) pathway and glucose metabolism (Fig. 2D). The VT Turquoise module showed overrepresentation for transcripts involved in extracellular matrix interactions and related signaling pathways (FDR<0.04) (Fig. 2D).

In DB tissues, 6 subnetworks of correlated expression were recognized (Fig. 3, Table S5). The DB Mustard, DB Tomato, and DB Magenta modules were most highly correlated with birthweight, while the DB Brown module was the only DB subnetwork with a significant blood pressure correlation (Fig. 3C). The DB Mustard module (Fig. 3D) was enriched for expressed genes involved in growth factor signaling and nuclear envelope function (FDR<0.07., hypergeometric test). Transcripts of the DB Tomato module (Fig. 3D) featured protein-coding transcripts involved in histone methylation, processing of mRNA, and Notch signaling (FDR<0.06). The DB Magenta group was highly enriched (FDR<0.001) for genes involved in host immune response pathways (Fig. 3D). The DB Brown module included transcripts encoding proteins involved in the HIF and transforming growth factor-β pathways in addition to those involved in the renin-angiotensin-aldosterone response, but these enrichments were not statistically significant (FDR>0.1) (Fig. 3D). The full module enrichment analysis results are shown in Figs. S4&S5.

**Figure 3.**
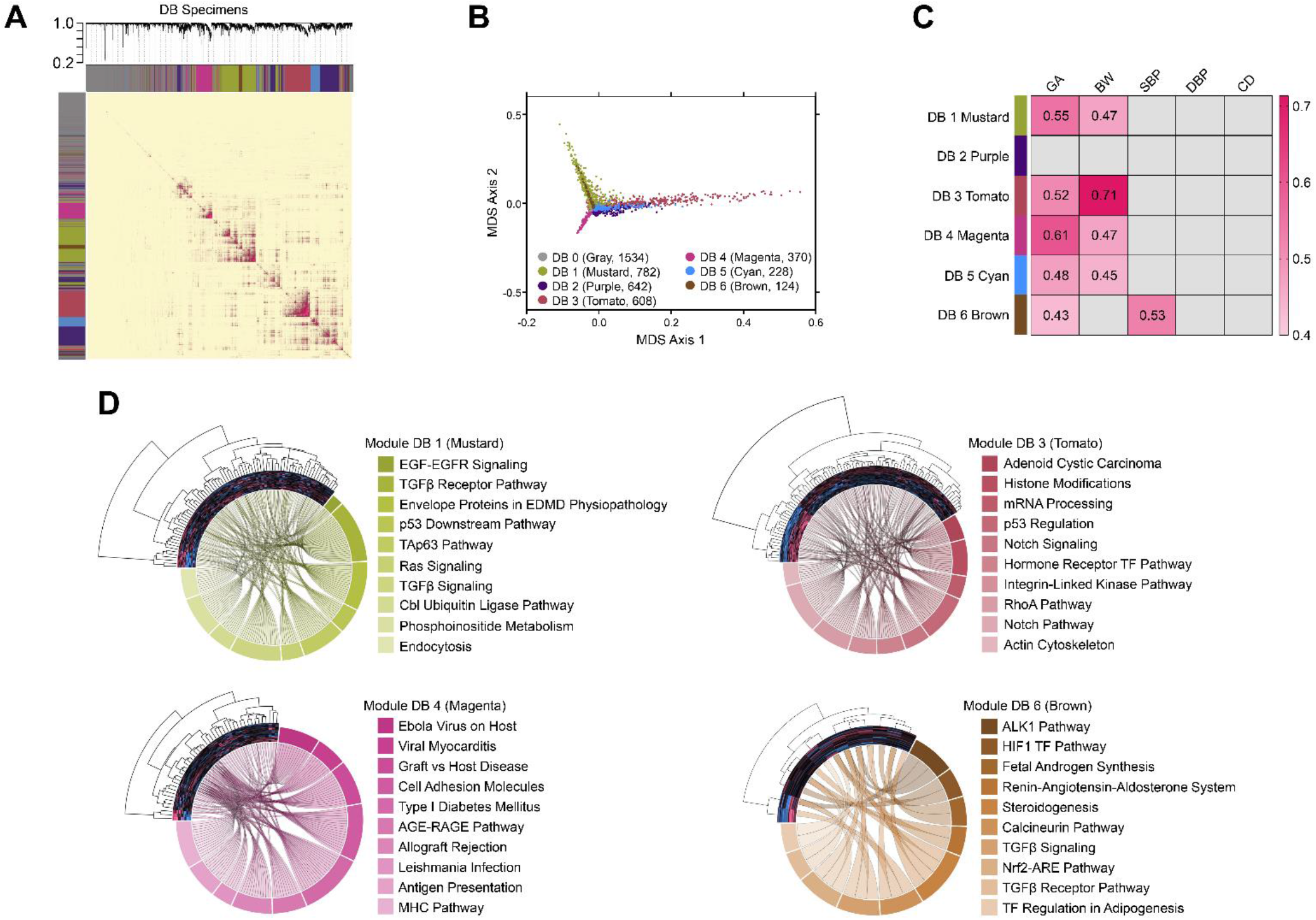
Consensus correlation network modules among decidua basalis (DB) placental specimens. WGCNA was applied to the most variable DB transcripts (top quartile, 4,288 RNAs). (A) Clustering dendrograms and topological overlap matrix heatmap (β=21, scale-free topology model fit r^2^ = 0.90, median connectivity=0.8). (B) Multimensional scaling scatterplot for the topological overlap matrix in panel A. (C) Module-trait correlation analysis results with statistically significant (*p*<0.05) correlation coefficients shown in colored boxes. (D) Chord diagrams depicting the top10 enriched pathways associated with select modules.

Modules within each anatomical placental region shared no common transcripts by algorithmic design. Between these regions, the degree of similarity among individual modules measured using the Jaccard index (JI) ranged from moderate (JI=17.2% between VT Orange and DB Tomato) to trivial (JI ≤3% for 30 module pairs) (Fig. S6).

### Co-expression Network Module Reproducibility and Predictive Performance Across Studies

Between the 2 structural placental regions (VT vs. DB), all transcript modules except for the DB Mustard group showed statistical evidence for presentation (Fig. 4 A&B). Given this, we proceeded to examine network module reproducibility more generally, using three previously published placental RNA-seq datasets in which similar clinical phenotypes were studied: 1) GSE148241 ^18^, consisting of EOPE (n=3), EOPE+FGR (n=6), and term control (n=32) specimens; 2) GSE114691 ^17^, comprising 79 samples including FGR (n=18), EOPE (n=20), EOPE+FGR (n=20), and PTB controls (n=21); and 3) PRJEB30656 ^19^ in which term FGR placental samples (n=5) were compared with term controls (n=5). For the GSE148241 dataset, the VT Blue, VT Black, and DB Brown modules were among those most strongly conserved, while the DB Tomato and DB Magenta modules were weakly preserved (Fig. S7 C&D). The module preservation patterns for the GSE114691 samples were like those of GSE148241, except for the weak conservation of the VT Fountain module (Fig. S7C&D). The PRJEB30656 placentas showed evidence for retention of 4 VT modules (Turquoise, Black, Straw, and Blue) and 3 DB modules (Mustard, Magenta, and Brown) (Fig. S7C&D).

**Figure 4.**
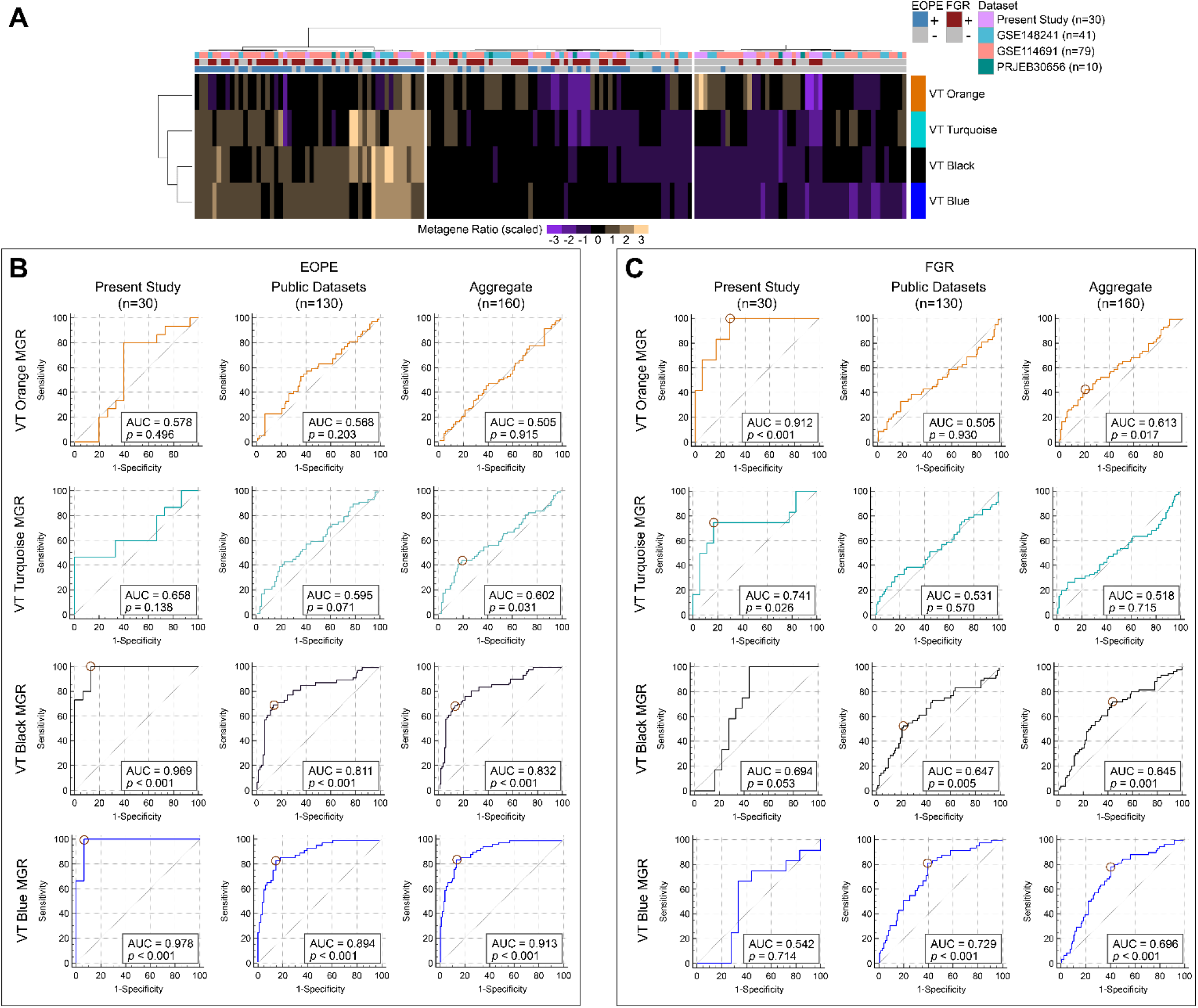
Metagene ratio (MGR) analysis for selected villous tissue (VT) correlation network modules and associations with clinical diagnoses. (A) Heatmap with unsupervised hierarchical clustering applied to MGRs for 4 VT co-expression network modules. (B, C) ROC curves for MGRs of selected VT modules in association with EOPE (B) and FGR (C). ROCs for the VT specimens in the present study (n=30), public datasets (n=130), and the aggregated set of all samples (n=160) are shown in the leftmost, middle, and rightmost columns of panels B and C, respectively. The points where Youden’s *J* is maximized (*J*_max_) are indicated by circles in cases where the AUC is significant (*p*<0.05).

Next, to gauge the utility of network modules in classifying clinical disease categories across studies (as may be applicable when detailed information on continuous clinical variables is inaccessible), we adapted and extended the metagene classification approach of Lauss and colleagues^9^. A metagene ratio (MGR) was calculated for each module, representing the quotient of the averaged expression of the transcripts most positively and negatively correlated with specific continuous clinical variables (Figs. S8&S9). These latter variables were selected based on their overall statistical association with each module in our specimens (Figs. 2C & 3C).

Cluster analysis applied to the VT module MGRs revealed 3 major groups among the 4 datasets: a set with elevated VT Black and VT Blue MGR expression comprising a majority of the EOPE samples; a cluster containing a large proportion of control specimens; and an intermediate group (Fig. 4A). By ROC analysis, the VT Black and VT Blue MGRs showed the highest predictive performance for EOPE but had more limited associations with FGR (Fig. 4 B&C). Although the VT Orange module was significantly correlated with birthweight (r=0.60, Fig. 2C) and had an MGR that was strongly associated with FGR in our samples (AUC=0.91, Fig. 4C), its association with FGR in the other datasets was weak (AUC=0.61, Fig. 4C). The VT Turquoise MGR showed only limited predictive ability for EOPE (Fig. 4B) and lacked consistently significant associations with FGR (Fig. 4C).

Among DB module MGRs, unsupervised clustering demonstrated 4 major categories: a group comprising mostly control specimens having elevated DB Tomato, DB Mustard, and DB Magenta MGRs with diminished DB Brown MGR levels; a small cluster with reduced expression of all DB module MGRs; and2 clusters containing a majority of the EOPE and FGR samples stratified by DB Brown module MGR expression (Fig. S10A). By ROC analysis, the DB Brown MGR was the most predictive for EOPE overall and exhibited some ability to detect FGR in the external datasets (Fig. S10B). Across studies, the MGRs of the DB Magenta, DB Mustard, and DB Tomato modules exhibited significant associations with FGR in our dataset but lacked consistent predictive performance more generally (Fig. S10C).

### Targeted Pathway Analysis Unveils a Reproducible Transcriptional Signature in a Subset of EOPE Placentas Suggestive of a Glycolytic Metabolic Shift

In reviewing our results, we noted that cluster analysis of the VT Black MGR transcripts subdivided the VT placental samples into distinct groups, including a set of EOPE±FGR samples with an expression pattern distinct from the remaining specimens (Figs. S8C & S11A). Since exploratory functional enrichment analysis for this module showed an overrepresentation of gene sets involved in metabolism (Fig. 2F), we performed a targeted analysis of the genes in the glycolysis/gluconeogenesis (KEGG hsa00010) pathway. These transcripts reproduced the clustering pattern observed for the VT Black MGR transcripts, with a subgroup sharing an increased abundance of genes encoding key glycolytic enzymes (callout in Fig. S11B). Consistent with the observed enrichment for hypoxia-responsive pathways in the VT Black module, this expression pattern suggested a bioenergetic shift away from aerobic oxidative phosphorylation towards increased lactate production ^22^.

To further simplify our analysis, we identified 2 RNA-seq expression ratios of VT Black module genes: *LDHA*/*LDHB*; and *PDK1*/*GOT1*. <u>T</u>he *LDHA* gene product preferentially converts pyruvate to lactate, whereas the *LDHB* gene product predominantly catalyzes the reverse reaction ^20^, and the expression ratio of the 2 has been used as a surrogate index for oxidative phosphorylation (Warburg effect) usage in cancer cells ^21^. The transcript for *PDK1* is a direct target of HIF-1α and a central regulator of the pyruvate dehydrogenase complex, a “gatekeeper” in hypoxia-related metabolic reprogramming ^22^. We prioritized *GOT1* because it is repressed by HIF-1α experimentally ^23^ and displayed expression reciprocal to *PDK1* in our dataset (r=-0.76, *p*<0.0001). Clustering based on these ratios reiterated the distinct subgroup within EOPE±FGR samples seen with the KEGG pathway transcripts (Fig. S11C). In the EOPE VT specimens, PC analysis of the VT Black MGR and KEGG hsa0010 pathway transcripts revealed 2 distinct sample clusters (Fig. S11D) strongly correlated with the *LDHA*/*LDHB* and *PDK1*/*GOT1* ratios (Fig. S11E). Division of the EOPE specimens into these 2 subgroups produced an expression pattern consistent with a glycolytic metabolic shift toward increased lactate production (Cluster 1) and a pattern suggestive of normoxic glucose utilization (Cluster 2) (Fig. S11F).The expression ratios in VT specimens by RNA-seq were significantly correlated with the corresponding ratios detected using qPCR (r=0.70, p<0.001 for *LDHA*/*LDHB*; r=0.56, *p*<0.002 for *PDK1*/*GOT1*; Fig. S12). A comparison of the RNA-seq expression ratios within and between the VT and DB anatomical placental regions for the paired samples (n=27) is shown in Fig. S13 where, in addition to the main VT cluster (Cluster 1), a second cluster of four samples exhibiting selectively elevated DB ratios was also noted (Fig. S13D).

We broadened our analysis to the 3 previously published datasets to determine whether these findings were reproducible. Clustering of the batch-corrected, normalized expression data for the VT Black MGR and KEGG hsa0010 transcripts consistently partitioned a common subset of samples (tan dendrogram lines, Fig. 5 A&B). This cluster could be reproduced using the RNA-seq expression ratios *LDHA*/*LDHB* and *PDK1*/*GOT1*, and included the Cluster 1 VT samples from our study (Fig. 5C). Across EOPE±FGR specimens from the available studies, combined analysis of these strongly correlated indices again revealed metabolic gene expression patterns consistent with a spectrum ranging from a standard bioenergetic profile to a shift similar to that seen in anaerobic glycolysis (Fig. 5 E&F).

**Figure 5.**
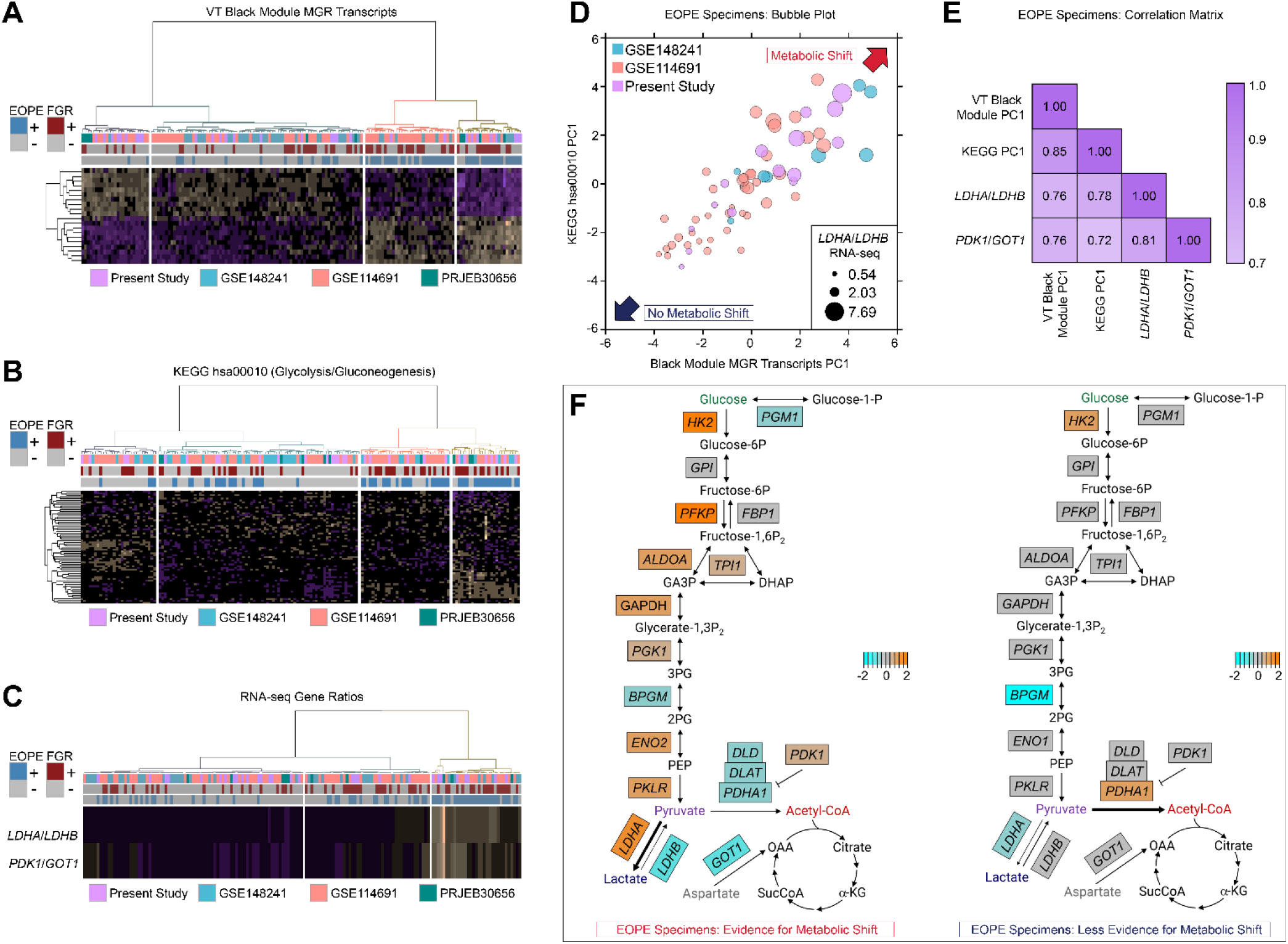
A reproducible molecular signature consistent with a glycolytic metabolic shift is observed in a subset of EOPE placenta specimens. (A-C) Heatmaps with unsupervised hierarchical clustering in samples from 4 studies (n=160) based on: (A) VT Black module MGR transcripts; (B) transcripts associated with the KEGG pathway hsa00010; and (C) normalized RNA-seq expression ratios of *LDHA*/*LDHB* and *PDK1*/*GOT1*. (D) Bubble plot comparing the first PCs of EOPE±FGR specimens , stratified by the expression VT Black module MGR and hsa00010 KEGG pathway transcripts. Bubble size correlates with the *LDHA*/*LDHB* RNA-seq expression ratios. (E) Pearson correlation matrix for pairs comprising PC1 values for VT Black MGR transcripts, PC1 values for KEGG hsa00010 transcripts, and the RNA-seq expression ratios for *LDHA*/*LDHB* and *PDK1*/*GOT1* (all *p*<0.01). (F) Pathway diagrams adapted from the KEGG hsa00010 map showing the expression of EOPE specimen transcripts, subdivided into metabolic subphenotypes based on transcriptional profiling evidence, relative to control samples.

## Discussion

In this placental transcriptional profiling study, WGNCA revealed 14 correlated gene modules in our dataset. All but 2 exhibited significant correlations with clinical variables (maternal blood pressure, birthweight, and gestational age at delivery) in our samples, providing a complementary alternative to differential abundance analysis for relating the placental transcriptome to EOPE and FGR. In considering the 160 samples from all 4 datasets, integrated analysis revealed 2 modules with strongly conserved EOPE and FGR associations: VT Blue (1001 transcripts, highly correlated to BP and enriched for membrane receptor signaling pathways), and DB Brown (124 transcripts, highly correlated to BP, comprised of transcripts involved in the regulation of the renin-angiotensin-aldosterone system, hypoxia-inducible factor, and steroidogenesis). These modules were 9.6% similar (by Jaccard index) and shared 99 genes in common, including several transcripts canonically associated with PE, such as *LEP* and *FLT1* ^24^. Of the 5 modules most strongly correlated with birthweight, 4 (VT Orange, DB Tomato, DB Magenta, and DB Mustard) were significantly associated with FGR in our specimens but lacked strong associations when the 4 datasets were considered in aggregate. Our identification of the VT Black module was of particular interest, given its independence from the larger VT Blue cluster, its strong association with EOPE across studies, and its functional enrichment for transcripts associated with HIF signaling and core cellular metabolism; we therefore focused further consideration around this co-expression subnetwork.

Throughout pregnancy, the placenta must continually support the developing fetus while simultaneously executing its own energy-intensive biosynthetic activities; as such, it has a particularly high metabolic activity and requirement for oxygen ^25^. Glucose is the main carbohydrate used by the uteroplacental system, and glycolysis serves as the central hub for placental metabolism ^25^. Under stress conditions, particularly when oxygen supply is reduced, the placenta must compensatively reprogram its metabolism to keep pace with bioenergetic demands. In general, such circumstances require a switch to less energetically efficient pathways (e.g., anaerobic glycolysis) than would be used otherwise. We thus anticipated that the placental insufficiency associated with EOPE and FGR — of which oxidative stress and uteroplacental hypoxia are cardinal features ^6^ — would involve the coordinated expression changes in the VT Black module transcripts. However, we were surprised by the degree of variability in this expression signature among samples, both in our dataset and in the EOPE specimens from public sources.

Closer examination of the glycolysis/gluconeogenesis pathway genes revealed a distinct molecular subgroup with a transcriptional signature consistent with a glycolytic shift; this cluster could be recapitulated using just the expression ratios of *LDHA*/*LDHB* and *PDK1*/*GOT1* (Fig. 5). Strikingly, roughly half of all EOPE specimens (with or without FGR) and most of the isolated FGR specimens fell outside of this molecular cluster. To contextualize our present findings in light of the molecular classifications put forth by Cox and collaborators ^11, 12^, we evaluated our data in comparison to the gene panels used by these investigators (Fig. S14). We approximated their “canonical” and “immunologic” subclasses by hierarchical clustering based on the relative expression of *TAP1* to *FLT1* and *LIMCH1*.

Although an imperfect surrogate to their qPCR-based classification strategy, our analysis suggests that the metabolic shift signature occurred in both molecular subtypes described by Cox and colleagues and could represent an independent source of information within this framework.

It is now recognized that the plasticity associated with cancer cell metabolism is more nuanced than originally suggested by the classical model of Warburg-type glucose metabolism ^22^. This is likewise the case with the intricacies of intermediary metabolism in the human placenta ^25^, and the current results should be interpreted with these particularities in mind. It has recently been proposed that placental metabolic programming (e.g., increased lactate production from glucose) may actually reflect a dynamic and adaptive physiological state, and that failure of such adaptations might contribute to the pathobiology of PE ^25^. While we cannot yet characterize our observations in terms of adaptation, decompensation, or malplacentation, it is possible that the altered glycolytic transcriptional patterns we identified may be transient, either spatially (through regional variation within the placenta) or temporally (e.g., in response to acute stressors). Further work using animal models ^26^ and *in vivo* functional imaging of the human placenta ^27^ might help clarify the extent and nature of dynamic metabolic changes in complicated pregnancies.

Among the FGR cases, several co-expression modules were significantly associated with birthweight and FGR in our samples. However, these modules showed inconsistent evidence for preservation and poor associations with FGR in the other cohorts in which this condition was explicitly studied (GSE114691 and PRJEB30656). Indeed, we could identify no characteristic transcriptional signatures uniting these three studies. Research into FGR pathobiology is challenging given the diverse etiologies that might contribute to compromised fetal growth ^28^. The clinical criteria used to diagnose FGR are continually evolving, and the distinction between constitutionally small for gestation age fetuses (SGA) and those with FGR is not always clear. Therefore, it is unsurprising that prior placental transcriptional profiling studies of FGR have reflected this heterogeneity, particularly given that the criterion of SGA alone has been frequently applied as a surrogate for FGR ^24^. While previous studies have reported overlap in the molecular processes between normotensive FGR and FGR associated with PE ^12, 19, 29^, a recent large and comprehensive study reported multiple instances of divergence between FGR and EOPE expression patterns for several long RNA classes ^30^.

Since the isolated FGR cases were delivered preterm for fetal indications, it is impossible to predict whether the trajectory of these pregnancies might have included PE manifestations. Further, we cannot exclude the possibility that a small number of the FGR cases harbored associated (and potentially causal) chromosomal aberrations that were undetectable by traditional karyotyping. Further research to increase insight into the placental dysfunction associated with growth-compromised fetuses may require emphasis on pathway-level dysregulation rather than focusing on individual gene expression patterns.

## Strengths and Limitations

Strengths of the present study include the availability of rich clinical phenotyping data for trait correlation analysis, integration with data from other studies to assess reproducibility, the separate consideration of VT and DB samples, and the use of gestational age-matched preterm controls in additional to term controls. The application of WGCNA as means for gene and preeclampsia sample classification has garnered increased research focus ^31–33^, and it will be of interest to see how this approach may be further used for molecular subphenotyping of this syndrome in the near future.

Our study has several limitations. First, we recognize that isolated tissue biopsies may not reflect the global functioning of the placenta. We attempted to mitigate this potential shortfall by extending our observations to studies in which samples were pooled from multiple placental regions, finding that our principal observations were upheld in these other cohorts. Second, in our design, we focused on EOPE and FGR using carefully selected specimens that we believed would represent cases with the greatest potential to benefit from further targeted research; however, this restriction did not allow us to explore other clinical manifestations of these conditions. Finally, while powerful, we recognize that the analytical methods applied also entail shortcomings ^7^. The foremost goal of this study was to explore the utility of correlation network analysis for prioritizing informative transcript modules in the settings of EOPE and FGR delivered before term. We recognize that identifying correlation networks more robustly would require a broader representation of the full clinical spectrum.

## Perspectives

Our results contribute to a growing body of literature demonstrating that PE and FGR are more heterogeneous conditions than can be appreciated by applying clinical diagnostic criteria alone. We provide novel evidence for a molecular subphenotype consistent with a glycolytic metabolic shift that occurs frequently, but not universally, in placental specimens, suggesting a spectrum of placental responses to these clinical conditions. Translating this work into actionable methods to inform targeted therapeutics and guide personalized care will require further integration with key clinical variables including minimally invasive screening of placenta-derived materials, such as microparticles or cell-free nucleic acids ^34^. Molecular subphenotyping through placental transcriptomics has promise to clarify and guide this work.

## Data Availability

The data that support the findings of this study are available from the corresponding author 
upon reasonable request. The RNA-seq data generated for this publication have been deposited in 
NCBI's Gene Expression Omnibus (GSE203507).

## Nonstandard Abbreviations and Acronyms

BW: birthweight
CD: cesarean delivery
DB: decidua basalis
DBP: diastolic blood pressure
EOPE: early-onset severe preeclampsia
FGR: fetal growth restriction
GA: gestational age of delivery
HELLP: hemolysis, elevated liver enzymes and low platelets
PTB: spontaneous idiopathic preterm birth
JI: Jaccard index
MGR: metagene ratio
PC: principal component
PE: preeclampsia
qPCR: quantitative reverse-transcriptase polymerase chain reaction
RNA-seq: RNA sequencing
RQ: relative quantification
SBP: ystolic blood pressure
TB: erm birth
VT: villous tissue
WGCNA: weighted gene correlation network analysis

## Author Contributions

Designed research: WEA, CSB, and IAB conceived and developed the study design. Performed research: CSB and IAB participated in patient recruitment, abstraction of clinical data, and the collection and processing of the biological specimens; WEA, GZ, and TLS executed the RNA extraction and qPCR experiments. Analyzed data: WEA, TLB, and IAB conducted the data analysis with input from CSB. Wrote the paper: WEA wrote the first draft of the manuscript with input from CSB, TLB, and IAB. All authors participated with reviewing the findings of the work and contributed with critical revisions to the drafts of the manuscript.

## Acknowledgments

This work was conducted using the high-performance computing resources of the University of Illinois at Chicago (Advanced Cyberinfrastructure for Education and Research). The authors gratefully acknowledge the Nucleic Acid Shared Resource at The Ohio State University Comprehensive Cancer Center for technical support and helpful discussions while this work was ongoing. The graphical abstract was created using BioRender.com. This first author would like to dedicate the manuscript to his father, the late William E. Ackerman III, MD, for inspiration in the pursuit of academic medicine.

## Novelty and Relevance

### What Is New?

This study demonstrates:

- A subphenotype consistent with a glycolytic metabolic shift occurs more frequently but not universally in placentas from pregnancies complicated by preeclampsia.
- The ratios of *LDHA*/*LDHB* and *PDK1*/*GOT1* mRNA transcripts may serve as surrogate indices for larger gene panels to identify metabolically stressed placentas.

### What Is Relevant?

- The 2 placental transcript ratios described in this study may add additional precision to the molecular subphenotyping of the placenta beyond want could be inferred from clinical characteristics alone.

### Clinical/Pathophysiological Implications?

- Our findings add to general knowledge that preeclampsia is a heterogeneous hypertensive syndrome and this heterogeneity extends to the placenta.

## Supplemental Material

### 1a. Detailed Methods

See also: Major Resources Table.

#### Recruitment and Tissue Collection

Transcriptomics analysis was performed using placentas from 30 pregnant individuals carefully phenotyped and categorized into the following clinical groups: (1) severe early-onset PE (EOPE) with appropriate fetal growth for gestational age (GA) (n=8); (2) EOPE with FGR (EOPE+FGR, n=7); (3) normotensive, nonanomalous preterm births (PTBs) complicated by early-onset fetal growth restriction (FGR, n=5); (4) spontaneous idiopathic PTB without FGR (PTB, n=5); and (5) uncomplicated pregnancies delivered at term in the absence of labor by cesarean section (TB, n=5). The specimens comprising the TB and PTB comparator groups have been reported previously ^1–3^, but were reevaluated starting from the raw data using the methodologies described herein. A summary of these samples is presented in Table S1.

This study defined PTB as the spontaneous onset of labor with delivery before 37 weeks GA. For all cases of PTB, triple I (intraamniotic infection and/or inflammation) was excluded based on negative results of amniotic fluid biochemical tests, negative bacterial cultures, and the absence of histologic chorioamnionitis or funisitis on placental examination ^1^.

FGR was diagnosed in the antepartum period and defined ultrasonographically as an estimated fetal growth <10th percentile for GA and assigned sex with abnormal umbilical artery Doppler velocimetry; the diagnosis was confirmed following delivery. A diagnosis of nonanomalous, idiopathic FGR was established in infants without detectable structural anomalies when pathological examination of the placenta and clinical work-up to rule out karyotype abnormalities did not provide a cause for growth restriction.

The clinical definition of PE was based on the criteria put forth by American College of Obstetricians and Gynecologists’ Task Force on Hypertension in Pregnancy. Only patients diagnosed before 34 weeks GA (early-onset) were included in this study. Severe EOPE was diagnosed clinically by one or more of the following criteria: (1) systolic blood pressure of >160 mm Hg or diastolic >110 mm Hg on at least 2 occasions 6 h apart; (2) >5 g protein excretion per 24 h urine collection, and/or persistent +3 proteinuria on dipstick testing; (3) oliguria <500 ml/24 h; (4) cerebral or visual disturbances; (5) pulmonary edema or cyanosis; (6) epigastric or right upper quadrant pain; (7) impaired liver function; and/or (8) thrombocytopenia; All patients with EOPE met at least 3 criteria for severe status.

Individuals were delivered by cesarean section for medical indications including breech presentation, non-reassuring fetal status remote from delivery, worsening pregnancy status, and/or suspected abruption (see Table S2). At enrollment, pregnancies with multifetal gestations, chromosomal aneuploidies, fetal structural anomalies, infection, and known comorbidities (e.g., diabetes, thrombophilias, thyroid disease, etc.) were excluded. In all cases, GA was established based on last menstrual period and/or ultrasonographic examination prior to 20 weeks of gestation. Clinical indications for ultrasound-guided amniocentesis to evaluate for intraamniotic infection and/or inflammation included symptoms of preterm labor, preterm premature rupture of membranes, advanced cervical dilatation (≥3 cm), and/or uterine contractions unresponsive to tocolysis.

#### Total RNA Extraction

Villous tissue (VT) and basal plate decidua basalis (DB) were collected as previously described^1^. The DB was separated from the VT immediately after delivery, and the individual matched specimens were flash-frozen in liquid nitrogen and stored at -80°C until processing. Total RNA was isolated using TRIzol reagent (Life Technologies, Grand Island, NY), including phenol-chloroform extraction, isopropanol precipitation, and ethanol wash steps ^1, 4, 5^. In all cases, RNA integrity was confirmed through visualization of intact 28S and 18S rRNA bands by agarose gel electrophoresis in the presence of Protector RNase Inhibitor (Roche, Indianapolis, IN). Prior to sequencing, RNA extracts were purified using the RNA Clean-Up and Concentration Kit (Norgen Biotek, Thorold, Ontario, Canada).

#### Bulk RNA Sequencing (RNA-seq) and Differential Abundance Analysis

RNA-seq libraries were constructed as described in prior reports ^1, 4, 5^ using the TruSeq Stranded Total RNA Sample Prep Kit with Ribo-Zero Gold (Illumina, San Diego, CA). The libraries were quality-controlled for appropriate mass using the Qubit 2.0 fluorometer (Life Technologies) and insert size using the BioAnalyzer 2100 system (Agilent, Santa Clara, CA). All sequenced samples had an RNA integrity number ≥6. Sequencing was performed using the Illumina HiSeq 2500 system and HiSeq version 3 sequencing reagents to generate 50 bp single-end reads. Clonal clusters were created using the Illumina cBOT platform with Illumina HiSeq version 3 cluster generation reagents to achieve a target density of approximately 800,000/mm^2^ per flow cell channel. Image analysis, base calling, and error estimation were performed using Illumina Analysis Pipeline in the HiSeq Control Software v2.2.38.

FastQC v0.11.8 was used for quality assessment of all sequenced reads. Reads were trimmed for quality (a minimum Phred quality score of 15) and contaminating adapter sequences with Trimmomatic v0.39 software, followed by mapping to the hg38 genome assembly using the TopHat2 v2.0.14 splice-aware alignment algorithm. Annotated feature counts were generated using HTseq v0.11.2. Sample scripts and code parameters used were as follows:

~~~
$ *fastqc -o qc <sample*_*name>.fastq.gz*
$ *for f in *.gz; do array=(${f//.fastq.gz/ }); java -jar*
      ${TRIMPATH}/trimmomatic-0.39.jar SE -threads 4 -phred33 $f
      *${OUTPATH}/${array*}_*trimmed.fastq
      ILLUMINACLIP:${ADAPTPATH}/TruSeq3-SE.fa:2:30:10 LEADING:3
      TRAILING:3 SLIDINGWINDOW:4:15 MINLEN:36; done*
$ *tophat2 -p 24 -a 6 -o ${OUTPATH}/<SAMPLE_NAME< --library-type*
      fr-firststrand ${BT2INDEX}/genome <SAMPLE_NAME<_trimmed.fastq
$ *for f in */accepted_hits.bam; do parentdir=’dirname $f’; parentdirname=’basename $parentdir’; samtools view $f*
      *| htseq-count --stranded=reverse - ${GTFFILE} >*
      ${OUTDIR}/${parentdirname}.count; done
~~~

Human Genome Organisation Gene Nomenclature Committee (HGNC) symbols were updated using HGNChelper v0.8.1, and additional metadata was assigned using AnnotationDbi v1.60.0, both within the R environment. Differential transcript abundance was determined using the DESeq2 v1.32.0 statistical package in R, applying multivariate negative binomial generalized linear models. Surrogate variable analysis, performed using the sva v3.40.0 library, was included to model unrecognized heterogeneity not otherwise accounted for in clinical variables. Significant changes in gene expression were based on raw *p*-values <0.05 and a false discovery rate (FDR) <0.1 to adjust for multiple testing. Statistical models included clinical outcome, assigned fetal sex at delivery, and the first two components of the surrogate variable analysis as covariates.

To minimize batch effects, the RNA-seq data reported in this report were generated in two rounds of sequencing. The PTB and TB specimens, reported on previously ^1–3^, were isolated and sequenced concurrently with samples S16, S20, S21, and S22 (Table S2). The original RNA-seq data generated for this publication have been deposited in NCBI’s Gene Expression Omnibus (GEO) and are accessible through accession number GSE203507.

#### Weighted Gene Correlation Network Analysis (WGCNA) and Module Preservation

Correlation network analysis was performed using the top quartile (∼4000) of normalized transcript feature counts exhibiting the greatest variability across samples in each placental region (VT and DB), as determined using standard error of the mean (SEM). Co-expression networks were assembled using the WGCNA v1.72-1 R library. First, similarity matrices were obtained using Pearson correlation coefficients (PCCs) between transcript pairs from log_2_-transformed gene expression data. Next, weighted adjacency matrices (encoding the connection strengths between pairs of nodes) were obtained by raising individual entries in a given similarity matrix to a power parameter β (i.e., node adjacency = |PCC|^β^), selected using the pickSoftThreshold() function to approximate a scale-free network topology. Topological overlap matrices (TOMs, unsigned type, with similarity measures reflective of the relative interconnectedness among individual nodes) were then calculated using the TOMsimilarity() function, together with the corresponding dissimilarity matrices (1-TOM). Finally, co-expression modules of highly interconnected transcripts were identified using an adaptive branch pruning approach by applying the Dynamic Tree Cut algorithm implemented with dynamicTreeCut v1.63-1.

Statistical evaluation for the preservation (robustness) of network modules identified by WGCNA was performed using the modulePreservation() function in the WGCNA R package. Summary *Z* statistics and *p*-values were calculated using log_2_-scaled, normalized gene expression data. The *Z*_summary_ (*Z*_s_) statistic, representing the average of density and connectivity-based network module preservation statistics, was interpreted as follows: *Z*_s_>10, strong evidence of module preservation; 2<*Z*_s_<10, weak to moderate evidence for preservation; and *Z*_s_<2 minimal evidence of preservation.

#### Eigengene-Based Assessment of Module-Trait Associations

Module-trait associations for groups of transcripts were computed by taking the modulus of the PCC between the eigengenes for transcripts of interest and selected continuous clinical variables. The eigengene for each module was defined as the first principal component of the scaled expression matrix and calculated using singular value decomposition implemented by the moduleEigengenes() function in the WGCNA package. The corPvalueStudent() function in the WGCNA library was used to calculate *p*-values for all module-trait correlations except for the binary categorical variable of cesarean delivery (yes or no), which was evaluated using point-biserial correlation with the cor.test() function.

#### Metagene Ratio (MGR) Analysis

As a complement to the eigengene-based module-trait correlation analysis applicable for continuous phenotypic trait variables, we adopted a metagene-centered approach to evaluate the predictive utility of individual modules in the classification of categorical variables using conventional receiver-operating characteristic (ROC) analysis. In this context, metagenes were defined as the arithmetic mean of normalized, variance-stabilizing transformed feature counts for selected groups of transcripts, as per the general approach described by Lauss et al. ^6^. Transcript selection was guided by each module’s overall gene-trait correlation analysis results. First, PCCs relating the abundance of individual transcripts in each module to continuous variables of clinical relevance (birthweight and maternal blood pressure) were calculated, then ranked according to the strength of association (taking the arithmetic mean of PCCs in cases where modules showed significant correlations with multiple phenotypic indices). Next, metagenes were computed for groups of transcripts with the greatest positive (n=10) and negative (n=10) correlation rankings. Finally, for each module, a metagene ratio (MGR) was quantified by dividing the mean expression of the positively correlated metagene by that of the negatively correlated metagene. MGRs so derived were subsequently assessed for their association with EOPE and FGR using ROC analysis, employing area under the ROC curve (AUC) as the performance metric.

#### Comparison and Integration with Previously Published Datasets

To assess the consistency of our present findings with those of prior studies, we performed a literature search for bulk RNA-sequencing datasets of the human placenta in the clinical contexts of early-onset preeclampsia and/or fetal growth restriction, in addition to querying the data repositories such as GEO and the European Nucleotide Archive (ENA) directly. We selected for consideration studies for which raw data (fastq files) had been deposited to repositories with unrestricted access, and with clinical information sufficient to allow a reasonable estimation of the criteria used to define PE and FGR. We aimed to obtain a representative, but not exhaustive, set of samples for comparison.

We identified three relevant, public placenta-derived RNA-seq datasets for further analysis: (1) GSE114691 ^7^, comprising 79 samples (18 with preterm FGR [defined as estimated fetal weight <10^th^ percentile for GA with abnormal umbilical and uterine artery Doppler evaluations], 20 with EOPE, 20 with EOPE+FGR, and 21 with preterm birth in the absence of other complications used as control specimens) from Ontario, Canada; (2) GSE148241 ^8^, consisting of 41 samples (6 from participants with severe EOPE+FGR, 3 from patients with severe EOPE without FGR, and 32 from subjects delivered at term) from the Guangdong Province of China; and (3) PRJEB30656 ^9^, which consisted of 5 term placental specimens from subjects with FGR (based on ultrasonographic estimation of fetal weight <10^th^ percentile with abnormal cerebroplacental ratio or uterine artery pulsatility index measurements, or an estimated fetal weight <3^rd^ percentile with normal vascular indices), and 5 uncomplicated term control samples, all from Olsztyn, Poland.

The same analytical workflow described above was used to generate feature counts from the available raw fastq files, except that paired-end trimming and mapping algorithms were employed for the samples from the GSE148241 and PRJEB30656 datasets. In instances where feature counts from multiple studies were integrated for joint analyses, study-dependent batch effects were removed using the removeBatchEffect() function from the limma v3.54.1 R package. A summary of the characteristics of these studies in relation to the present study is presented in Table S3

#### Functional Enrichment Analysis

We performed functional enrichment on prioritized groups of transcripts using the clusterProfiler v4.6.0 R package, applying curated gene sets archived in the Molecular Signatures Database (MSigDB) Canonical Pathways v7.4 having sizes between 10 and 250 members, inclusive.

Overrepresentation was calculated using hypergeometric testing employed by the enricher() analyzer function. Chord diagrams for top enriched pathways (ranked according to *p*-value in ascending order) were rendered using the circlize v0.4.15 R library. Where indicated, gene expression datasets were intersected with curated gene lists from the Therapeutic Target Database (https://db.idrblab.net/ttd/), a repository of known and potential druggable targets, and the Comparative Toxicogenomics Database, a compendium of curated gene-disease associations (http://ctdbase.org/).

#### qPCR

Guided by our WGCNA results, we selected 4 transcripts, lactate dehydrogenase A (*LDHA*), lactate dehydrogenase B (*LDHB*), pyruvate dehydrogenase kinase 1 (*PDK1*), and glutamic-oxaloacetic transaminase 1 (*GOT1*) for cross-validation by qPCR using the general methodology described previously ^4, 5^. These transcripts were chosen based on their relevance to core placental metabolic pathways. For all reactions, reverse transcription was achieved using Superscript II Reverse Transcriptase (Invitrogen) with oligo(dT) primers. The following TaqMan gene expression assays (Thermo Fisher Scientific) were used: *LDHA*: Hs01378790_g1, *LDHB*: Hs00600794_mH, *PDK1*: Hs00176853_m1, *GOT1*: Hs00157798_m1.The geometric mean of the Ct values for β2-microglobulin (*B2M*, Hs99999907_m1) and ribosomal protein L30 (*RPL30*, Hs00265497_m1) was used as a reference in each reaction.

Every 20 μL reaction comprised 1 μL cDNA (500 ng), 1 μL of TaqMan Gene Expression Assay, 10 μL TaqMan Fast Advanced Master Mix (Thermo Fisher Scientific), and 8 μL of nuclease-free water. All reactions were performed in duplicate. The relative abundance of each mRNA was calculated using comparative Ct method. The transcript ratio *LDHA*/*LDHB* was calculated as 2^–ΔC^, with ΔC representing the difference between the Ct values for *LDHA* and *LDHB* (i.e., ΔC = Ct*^LDHA^* – Ct*^LDHB^*). The *PDK1*/*GOT1* ratio was calculated in a similar manner. Correlations between the expression levels from RNA-seq and qPCR experiments were examined using Pearson’s product-moment correlation (on log-transformed data).

#### Statistics

All statistical analyses were performed using a combination of functions and packages within the open-source R environment v4.1.0 (https://www.R-project.org/), in addition to Prism v9 (GraphPad Software, La Jolla, CA) and MedCalc v20.027 (MedCalc Software Ltd, Ostend, Belgium) statistical software. Cytoscape v3.9.0 (https://cytoscape.org/) was used for network graph manipulation.

Clinical characteristics of the study group were summarized as mean ± standard deviation (SD) or median and interquartile range (IQR) for continuous variables, and group percentages for categorical variables. Clinical characteristics among of the study groups were evaluated using (analysis of variance ANOVA) or Kruskal–Wallis testing (for continuous variables) and Chi squared tests (for categorical variables). For these comparisons, a *p*-value <0.05 was considered statistically significant. Adjustments for multiple comparisons were performed using the Benjamini-Hochberg procedure, and in these cases, FDR<0.1 was considered significant.

Distance matrix calculations and hierarchical clustering were achieved using the dist() and hclust() R functions, respectively. Principal component (PC) analysis was performed on consensus pools transcripts using the prcomp() function in the R stats library. Consensus transcripts were defined as unique transcripts for each placental region assembled by taking the union of statistically significant (FDR<0.1) RNAs present in at least on pairwise statistical comparison in multivariate models evaluated using DESeq2 software using models that included clinical outcome, assigned fetal sex at delivery, and the first two components of the surrogate variable analysis as covariates.

Classic multidimensional scaling (MDS) was applied to dissimilarity based on topological overlap using the cmdscale() function in the R stats package. Convex hull calculations were performed using convhulln() in the geometry v0.4.6.1 R package. ROC renderings were generated using MedCalcv20.027 (MedCalc Software Ltd). Calculation and statistical testing of correlations were performed using the cor) and cor.test() functions in the R stats library. The corrplot v0.92 library was used to assist with correlogram plotting. Heatmaps were generated using the pheatmap v1.0.12 and ComplexHeatmap v2.14.0 R packages.

### 1b. Supplemental Results

#### Differential Abundance Results in Pairwise Statistical Comparisons

Among the VT samples, the EOPE specimens exhibited the greatest dispersion along the PC coordinates and showed considerable overlap with the EOPE+FGR tissues (Fig. 1B). In the DB specimens, co-clustering was noted between PTB and TB tissues, with intermixing among the EOPE, EOPE+FGR, and FGR groups (Fig. 1D). Furthermore, the EOPE and FGR DB biopsies showed pronounced dispersal, with the latter having appreciably more scattering in the PC space compared to the corresponding FGR VT tissues (Fig. 1 B&D).

For the VT samples, there were 4,772 and 4,414 differentially abundant (FDR<0.1) transcripts in the FGR and EOPE+FGR VT biopsies, respectively, relative to the PTB VT specimens (Fig. S1). Although comparatively fewer (2,692) transcripts were significantly altered in the EOPE VT samples relative to the PTB VT group, this was consistent with the more dispersed clustering observed in PC analysis (Fig. 1B). Similar trends were noted when these specimens were compared with the TNL VT samples, with 4,330, 3,984, and 2,345 differentially abundant transcripts for the FGR, FGR+EOPE, and EOPE groups, respectively. The VT specimens from pregnancies complicated by FGR alone differed from the EOPE VT samples by 1,013 expressed genes, while the EOPE+FGR VT samples differed from the FGR and EOPE VT tissues by 99 and 38 RNA species, respectively. These results agreed with the overall similarity rankings shown in Fig. 1A.

For the DB specimens (Fig. S2), the FGR, EOPE+FGR, and EOPE samples differed from the PTB samples by 921, 876, and 1,095 transcripts, respectively. Relative to the TNL DB placenta specimens, the FGR, EOPE+FGR, and EOPE samples had 1,565, 1,443, and 1,819 differentially abundant transcripts, respectively. There were 834 differentially abundant RNAs when FGR samples were compared with EOPE, 235 when EOPE+FGR was contrasted with FGR, and 13 when EOPE was compared against EOPE+FGR, in accord with the hierarchy of similarity scores for these groups (Fig. 1C).

In pairwise comparisons between the placental subregions in each of the clinical conditions (EOPE, EOPE+FGR, and isolated FGR), the DB specimens consistently exhibited a greater abundance of transcripts enriched or specific to this region compared to the VT (Fig. S3), consistent with our previously reported findings ^1^. Nevertheless, relative enrichment for certain RNA species in the VT region was also noted (Fig. S3).

## 2. Supplemental Tables.

**Table S1.**
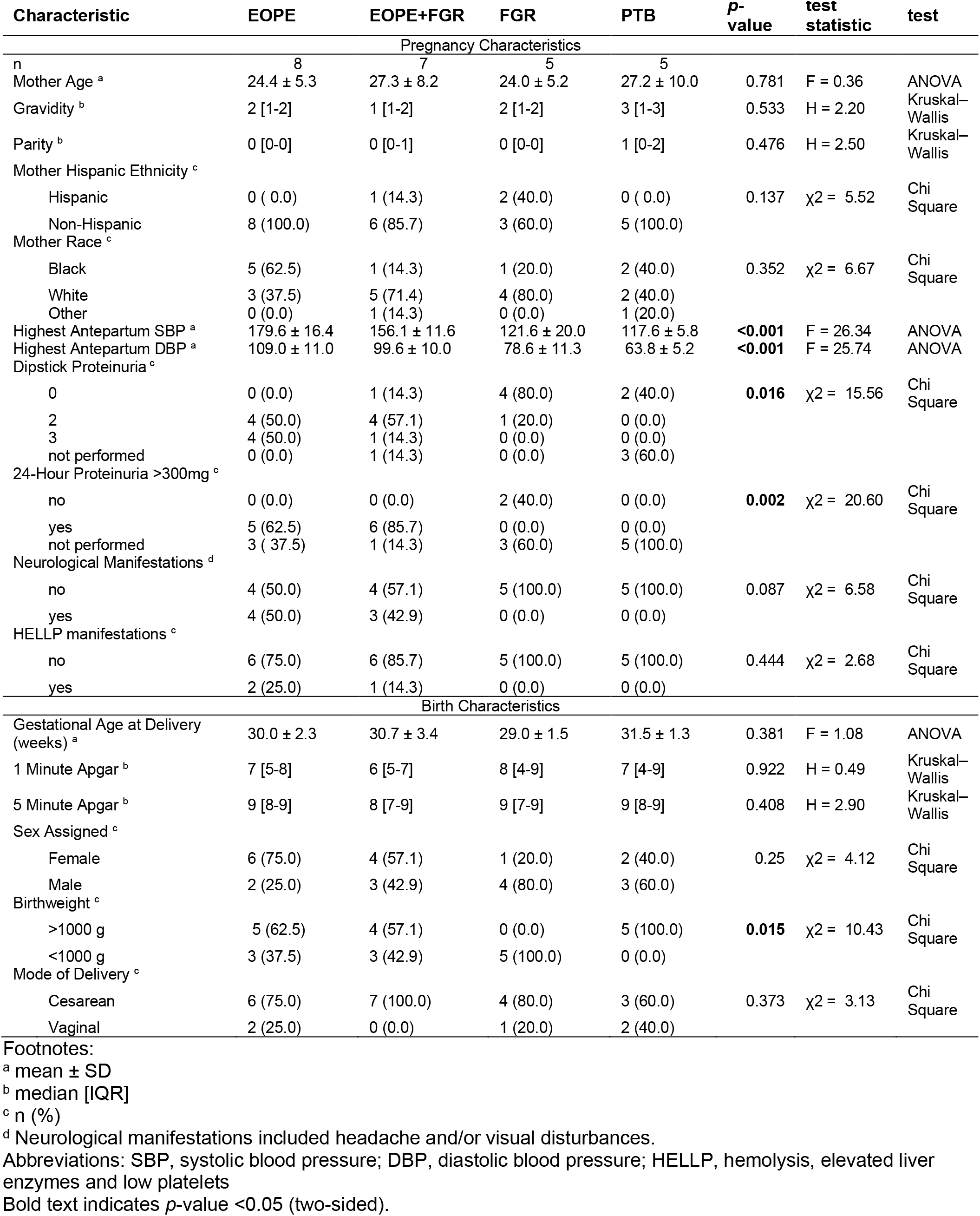
Summary of clinical characteristics for present study.

| Characteristic | EOPE | EOPE+FGR | FGR | PTB | p-value | test statistic | test |
| --- | --- | --- | --- | --- | --- | --- | --- |
| Pregnancy Characteristics |  |  |  |  |  |  |  |
| n | 8 | 7 | 5 | 5 |  |  |  |
| Mother Age <sup>a</sup> | 24.4 ± 5.3 | 27.3 ± 8.2 | 24.0 ± 5.2 | 27.2 ± 10.0 | 0.781 | F = 0.36 | ANOVA |
| Gravidity <sup>b</sup> | 2 [1-2] | 1 [1-2] | 2 [1-2] | 3 [1-3] | 0.533 | H = 2.20 | Kruskal-Wallis |
| Parity <sup>b</sup> | 0 [0-0] | 0 [0-1] | 0 [0-0] | 1 [0-2] | 0.476 | H = 2.50 | Kruskal-Wallis |
| Mother Hispanic Ethnicity <sup>c</sup> |  |  |  |  |  |  |  |
| Hispanic | 0 ( 0.0) | 1 (14.3) | 2 (40.0) | 0 ( 0.0) | 0.137 | χ<sup>2</sup> = 5.52 | Chi Square |
| Non-Hispanic | 8 (100.0) | 6 (85.7) | 3 (60.0) | 5 (100.0) |  |  |  |
| Mother Race <sup>c</sup> |  |  |  |  |  |  |  |
| Black | 5 (62.5) | 1 (14.3) | 1 (20.0) | 2 (40.0) | 0.352 | χ<sup>2</sup> = 6.67 | Chi Square |
| White | 3 (37.5) | 5 (71.4) | 4 (80.0) | 2 (40.0) |  |  |  |
| Other | 0 (0.0) | 1 (14.3) | 0 (0.0) | 1 (20.0) |  |  |  |
| Highest Antepartum SBP <sup>a</sup> | 179.6 ± 16.4 | 156.1 ± 11.6 | 121.6 ± 20.0 | 117.6 ± 5.8 | **<0.001** | F = 26.34 | ANOVA |
| Highest Antepartum DBP <sup>a</sup> | 109.0 ± 11.0 | 99.6 ± 10.0 | 78.6 ± 11.3 | 63.8 ± 5.2 | **<0.001** | F = 25.74 | ANOVA |
| Dipstick Proteinuria <sup>c</sup> |  |  |  |  |  |  |  |
| 0 | 0 (0.0) | 1 (14.3) | 4 (80.0) | 2 (40.0) | **0.016** | χ<sup>2</sup> = 15.56 | Chi Square |
| 2 | 4 (50.0) | 4 (57.1) | 1 (20.0) | 0 (0.0) |  |  |  |
| 3 | 4 (50.0) | 1 (14.3) | 0 (0.0) | 0 (0.0) |  |  |  |
| not performed | 0 (0.0) | 1 (14.3) | 0 (0.0) | 3 (60.0) |  |  |  |
| 24-Hour Proteinuria >300mg <sup>c</sup> |  |  |  |  |  |  |  |
| no | 0 (0.0) | 0 (0.0) | 2 (40.0) | 0 (0.0) | **0.002** | χ<sup>2</sup> = 20.60 | Chi Square |
| yes | 5 (62.5) | 6 (85.7) | 0 (0.0) | 0 (0.0) |  |  |  |
| not performed | 3 ( 37.5) | 1 (14.3) | 3 (60.0) | 5 (100.0) |  |  |  |
| Neurological Manifestations <sup>d</sup> |  |  |  |  |  |  |  |
| no | 4 (50.0) | 4 (57.1) | 5 (100.0) | 5 (100.0) | 0.087 | χ<sup>2</sup> = 6.58 | Chi Square |
| yes | 4 (50.0) | 3 (42.9) | 0 (0.0) | 0 (0.0) |  |  |  |
| HELLP manifestations <sup>c</sup> |  |  |  |  |  |  |  |
| no | 6 (75.0) | 6 (85.7) | 5 (100.0) | 5 (100.0) | 0.444 | χ<sup>2</sup> = 2.68 | Chi Square |
| yes | 2 (25.0) | 1 (14.3) | 0 (0.0) | 0 (0.0) |  |  |  |
| Birth Characteristics |  |  |  |  |  |  |  |
| Gestational Age at Delivery (weeks) <sup>a</sup> | 30.0 ± 2.3 | 30.7 ± 3.4 | 29.0 ± 1.5 | 31.5 ± 1.3 | 0.381 | F = 1.08 | ANOVA |
| 1 Minute Apgar <sup>b</sup> | 7 [5-8] | 6 [5-7] | 8 [4-9] | 7 [4-9] | 0.922 | H = 0.49 | Kruskal-Wallis |
| 5 Minute Apgar <sup>b</sup> | 9 [8-9] | 8 [7-9] | 9 [7-9] | 9 [8-9] | 0.408 | H = 2.90 | Kruskal-Wallis |
| Sex Assigned <sup>c</sup> |  |  |  |  |  |  |  |
| Female | 6 (75.0) | 4 (57.1) | 1 (20.0) | 2 (40.0) | 0.25 | χ<sup>2</sup> = 4.12 | Chi Square |
| Male | 2 (25.0) | 3 (42.9) | 4 (80.0) | 3 (60.0) |  |  |  |
| Birthweight <sup>c</sup> |  |  |  |  |  |  |  |
| >1000 g | 5 (62.5) | 4 (57.1) | 0 (0.0) | 5 (100.0) | **0.015** | χ<sup>2</sup> = 10.43 | Chi Square |
| <1000 g | 3 (37.5) | 3 (42.9) | 5 (100.0) | 0 (0.0) |  |  |  |
| Mode of Delivery <sup>c</sup> |  |  |  |  |  |  |  |
| Cesarean | 6 (75.0) | 7 (100.0) | 4 (80.0) | 3 (60.0) | 0.373 | χ<sup>2</sup> = 3.13 | Chi Square |
| Vaginal | 2 (25.0) | 0 (0.0) | 1 (20.0) | 2 (40.0) |  |  |  |

<sup>a</sup> mean ± SD<sup>b</sup> median [IQR]<sup>c</sup> n (%)<sup>d</sup> Neurological manifestations included headache and/or visual disturbances.
Abbreviations: SBP, systolic blood pressure; DBP, diastolic blood pressure; HELLP, hemolysis, elevated liver enzymes and low platelets
Bold text indicates p-value <0.05 (two-sided).

Table S2. Detailed clinical characteristics for present study.

Please see:

https://uofi.box.com/s/8kj1wac03sq9x4lw7148d8tyvbrcvm4q

Alternate repository:

https://osf.io/pa5wq/

doi; 10.17605/OSF.IO/PA5WQ

**Table S3.** Comparisons of study criteria, sampling methods, and sequencing methods among datasets considered.

| Study Cohort | Sample Size: n | GA at Delivery (mean±SD) | FGR Inclusion Criteria | PE Inclusion Criteria | Placental Sampling | Sequencing |
| --- | --- | --- | --- | --- | --- | --- |
| Present Study (GSE203507) | • EOPE: 8<br>• FGR: 5<br>• EOPE+FGR: 7<br>• PTB: 5<br>• TB: 5 | • EOPE: 29.8±2.3 wks<br>• FGR: 29.1±1.4 wks<br>• EOPE+FGR: 30.7±3.4 wks<br>• PTB: 31.6±1.9 wks<br>• TB: 39.0±0.1 wks | Ultrasonographic EFW < 10th percentile for GA and assigned sex with abnormal Doppler evaluations | ACOG Task Force on Hypertension in Pregnancy 2013 [PubMed: 24150027] | Single placental biopsy, separated into villous tissue and decidua basalis tissue | • Ribosome depletion<br>• 50 bp single-end<br>• Illumina HiSeq 2500 platform |
| GSE148241 | • EOPE: 3<br>• EOPE+FGR: 6<br>• Control: 32 | • EOPE: 30.0±2.0 wks<br>• EOPE+FGR: 32.9±1.1 wks<br>• Control: 38.8±2.3 wks | Not specified | ACOG Task Force on Hypertension in Pregnancy 2013 [PubMed: 24150027] | Villous tissues collected from mid-section of placenta between chorionic and basal surfaces at 4 different positions | • Poly-A enrichment<br>• 125 bp paired-end<br>• Illumina HiSeq 2500 platform |
| GSE114691 | • EOPE: 20<br>• FGR: 18<br>• EOPE+FGR: 20<br>• Control: 21 | < 34 weeks at diagnosis<br>• EOPE: 29.6±3.1 wks<br>• FGR: 32.3±3.4 wks<br>• EOPE+FGR: 29.4±2.5 wks<br>• Control: 30.6±2.6 wks | Ultrasonographic EFW < 10th percentile for GA and assigned sex with abnormal umbilical and uterine artery Doppler evaluations | Hypertension (blood pressure > 140/90 mmHg) and proteinuria (≥ 300 mg in 24 h) | Villous tissue pooled from 2 central and 2 peripheral placental regions | • RNA enrichment/depletion method not specified<br>• 50 bp single-end<br>• Illumina HiSeq 2000 platform |
| PRJEB30656 | • FGR: 5<br>• Control: 5 | Term | Ultrasonographic EFW < 10th percentile for GA with abnormal cerebroplacental or uterine artery Doppler evaluations, or normal Doppler evaluations with EFW ≤ 3rd percentile for GA | Not included | Placenta biopsy (details of placental sampling not specified) | • Poly-A enrichment<br>• 50 bp paired-end<br>• Illumina HiSeq 4000 platform |

**Abbreviations:**
ACOG, American College of Obstetricians and Gynecologists
EOPE, early-onset preeclampsia
FGR, fetal growth restriction
GA, gestational age
PTB, spontaneous idiopathic preterm birth
PE, preeclampsia
wks, weeks

Table S4. Transcripts assigned to co-expression modules by correlation network analysis in villous tissue specimens.

*Please see:* https://uofi.box.com/s/jfwzaord2fn7xzc6krk023ow1pamf6pa

Alternate repository: https://osf.io/pa5wq/

doi; 10.17605/OSF.IO/PA5WQ

Table S5. Transcripts assigned to co-expression modules by correlation network analysis in decidua basalis specimens.

*Please see:* https://uofi.box.com/s/x81ml7u8p1ltiollhmou7wean6a0t9ib

Alternate repository: https://osf.io/pa5wq/

doi: 10.17605/OSF.IO/PA5WQ

Note: Supplemental spreadsheets are available at: https://uofi.box.com/s/h1fraq80bydl8dvq9rui7x21rahjoi2r

(These supplemental spreadsheets will also be archived and maintained at UIC INDIGO, https://researchguides.uic.edu/indigo/about, hosted by the University of Illinois-Chicago University Library).

## 3. Supplemental Figures and Figure Legends

**Figure S1.**
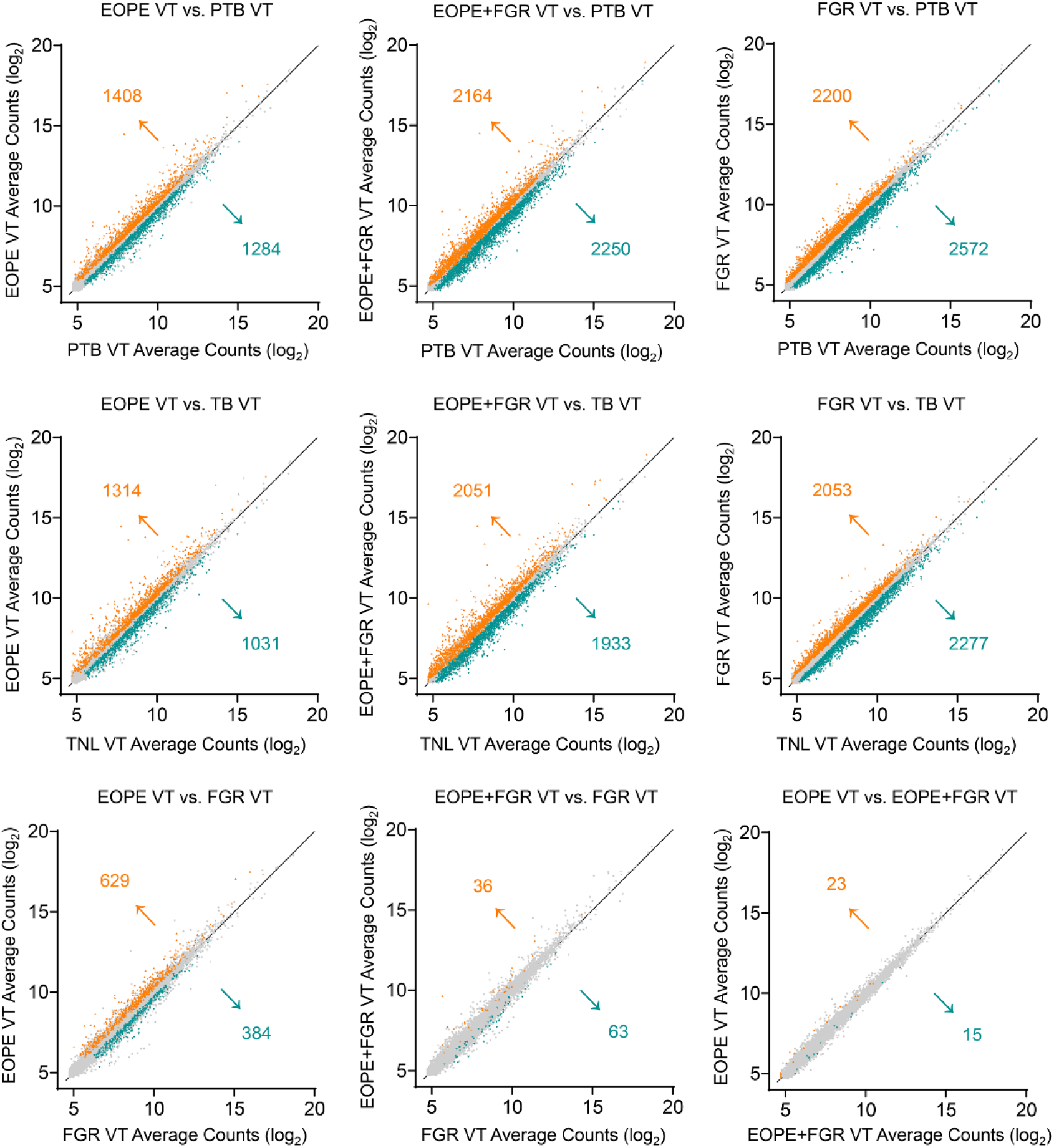
Overview of differential transcriptional abundance patterns in placental villous tissue (VT) samples. Scatterplots of average transcript expression data following variance stabilizing transformation with log_2_-scaling, showing the overall degree of correlation and differential abundance (FDR<0.1, colored triangles) in pairwise contrasts as determined using DESeq2 statistical software with surrogate variable analysis. The number of differentially abundant transcripts associated with each axis is shown adjacent to the arrows, which indicate the direction of the change. Gray dots represent RNA species that did not differ in abundance based on statistical models. Statistical models included clinical outcome, fetal sex, and the first two surrogate variable components as covariates. Abbreviations: EOPE, early-onset severe preeclampsia; FGR, fetal growth restriction; PTB, spontaneous idiopathic preterm birth; TB, term birth.

**Figure S2.**
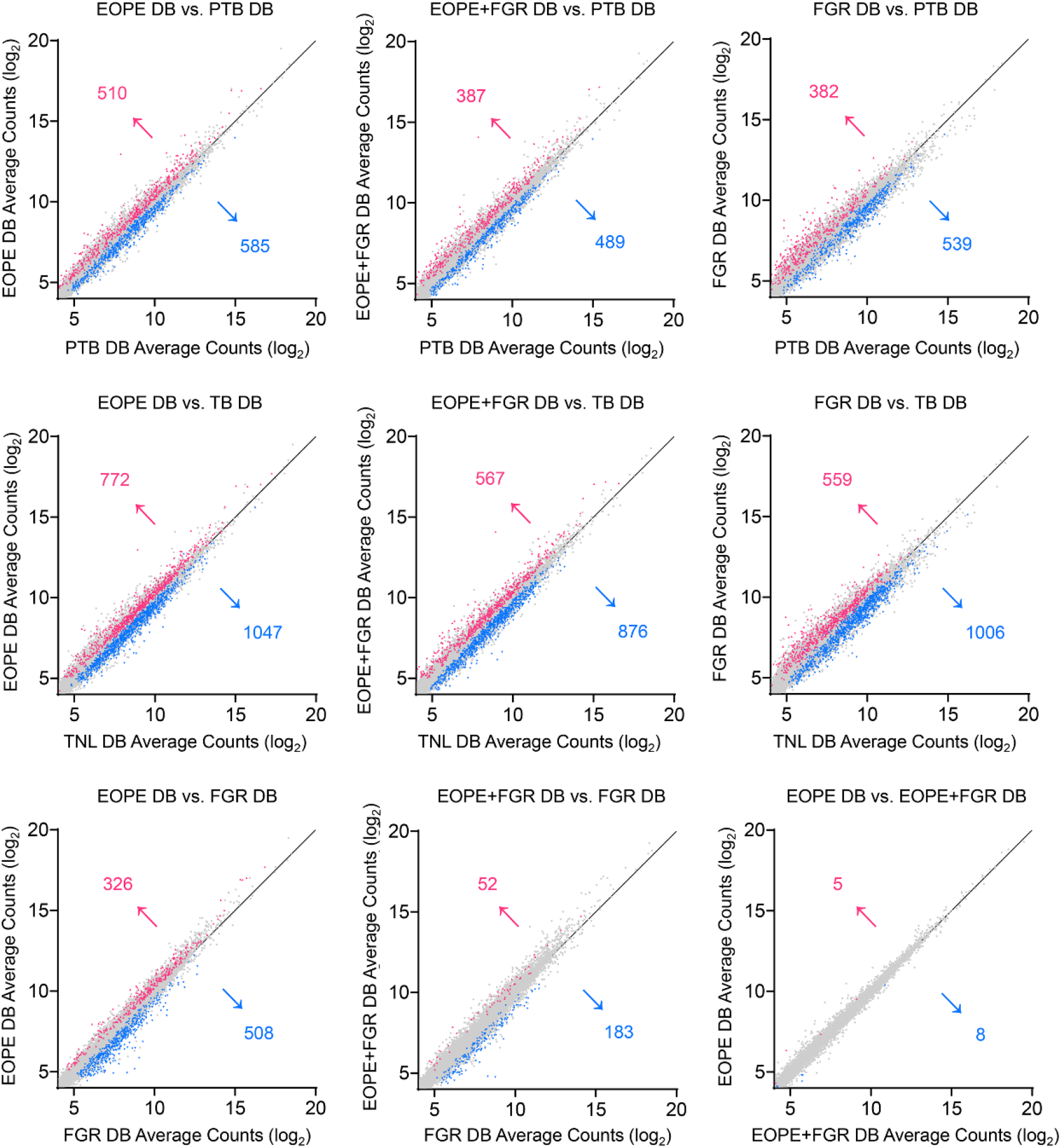
Overview of differential transcriptional abundance patterns in placental decidua basalis (DB) samples. Scatterplots of average transcript expression data following variance stabilizing transformation with log_2_-scaling, showing the overall degree of correlation and differential abundance (FDR<0.1, colored triangles) in pairwise contrasts as determined using DESeq2 statistical software with surrogate variable analysis. Arrows and numbers indicate the direction and amount of change toward either axis. Gray dots represent RNA species that did not differ in abundance based on statistical models. Statistical models included clinical outcome, feal sex, and the first two surrogate variable components as covariates. Abbreviations: EOPE, early-onset severe preeclampsia; FGR, fetal growth restriction; PTB, spontaneous idiopathic preterm birth; TB, term birth.

**Figure S3.**
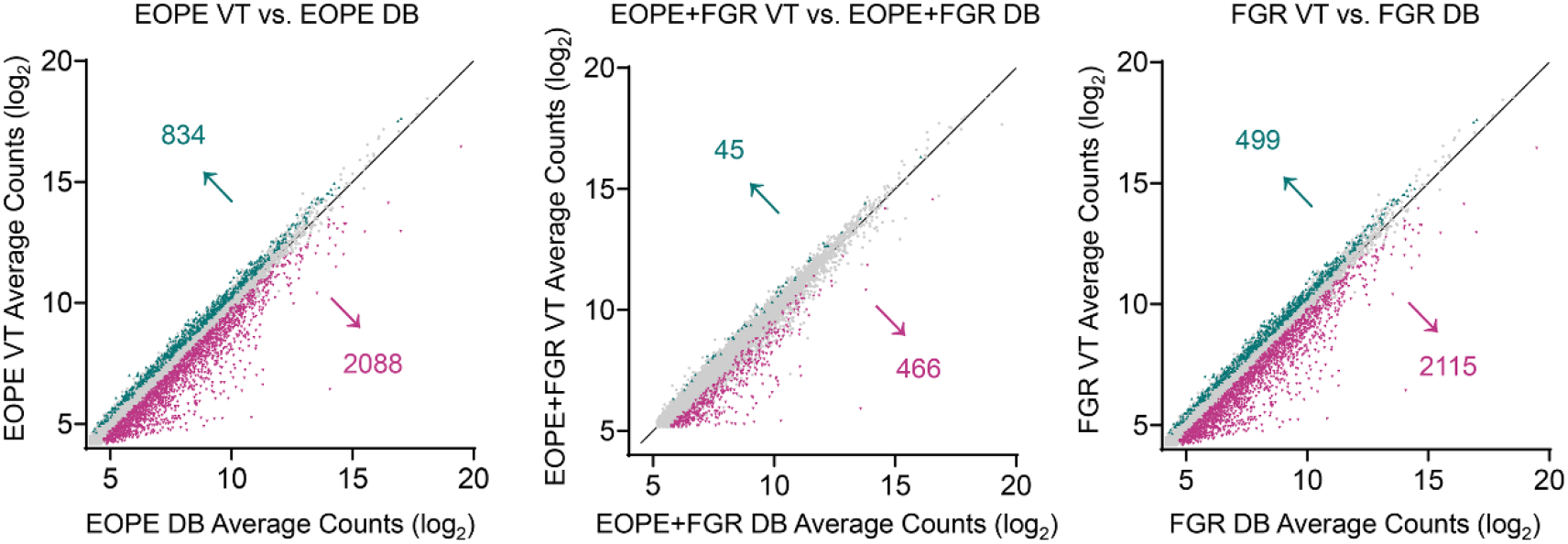
Overview of differential transcriptional abundance patterns in paired placental villous tissue (VT) and decidua basalis (DB) samples under each clinical condition. Scatterplots of the average transcript expression data after variance stabilizing transformation with log_2_-scaling, showing the overall degree of correlation and differential abundance (FDR<0.1, colored triangles) in pairwise contrasts determined using DESeq2 statistical software. The number of differentially abundant transcripts corresponding to each axis is shown adjacent to the arrows. Gray dots represent RNA species that did not differ in abundance based on pairwise statistical models. Abbreviations: EOPE, early-onset severe preeclampsia; FGR, fetal growth restriction.

**Figure S4.**
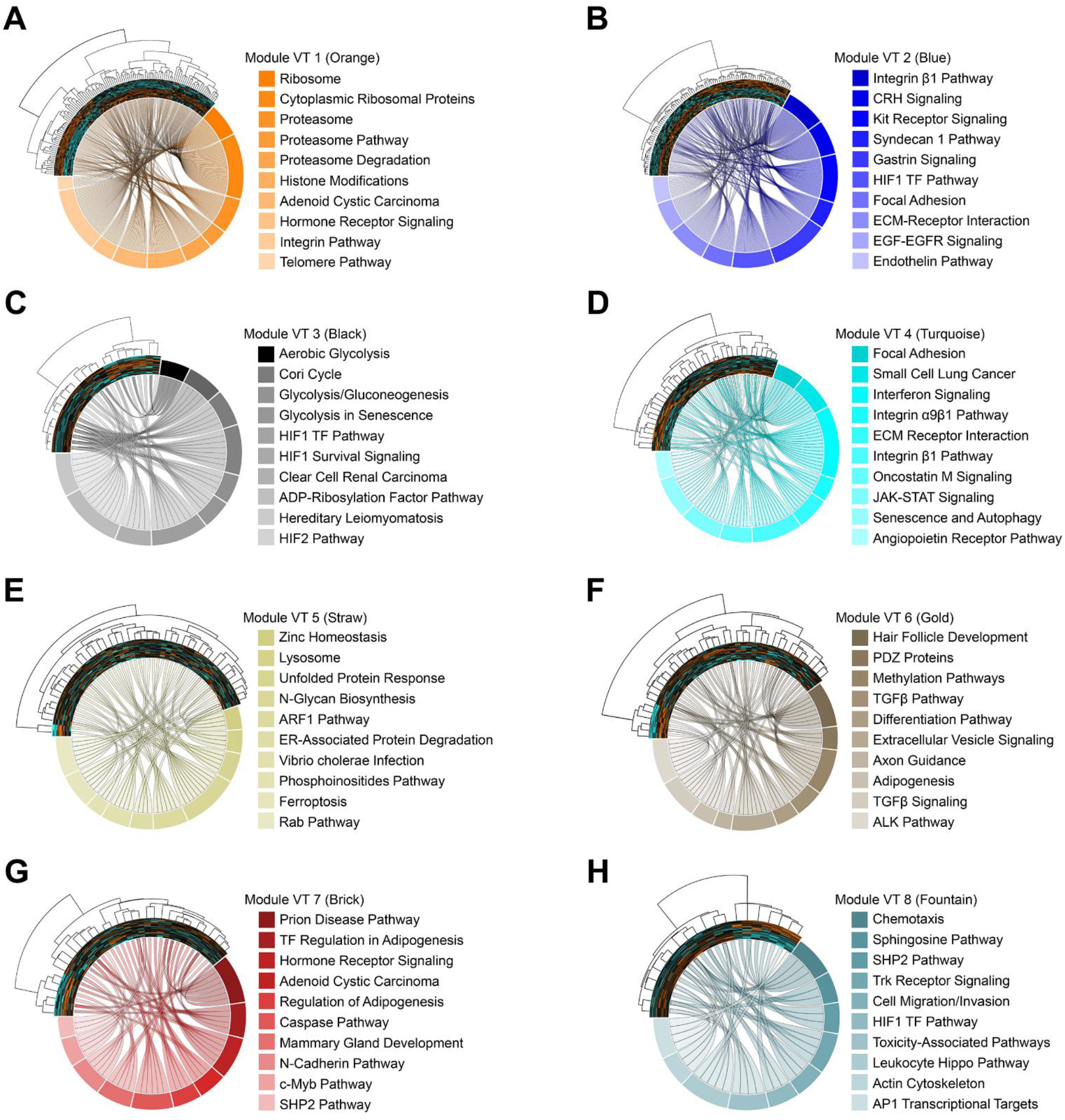
Pathway enrichment for all co-expression modules identified in the villous tissue (VT) samples. (A-H) Chord diagrams representing the top10 enriched pathways associated with the transcripts (depicted as clustered heatmaps) for the 8 VT network modules, ranked according to *p*- value. Note that the Gray Module (unassigned transcripts) was not included in this analysis. Each chord represents the connection between an individual module transcript in the heatmap and its presence in a given gene set.

**Figure S5.**
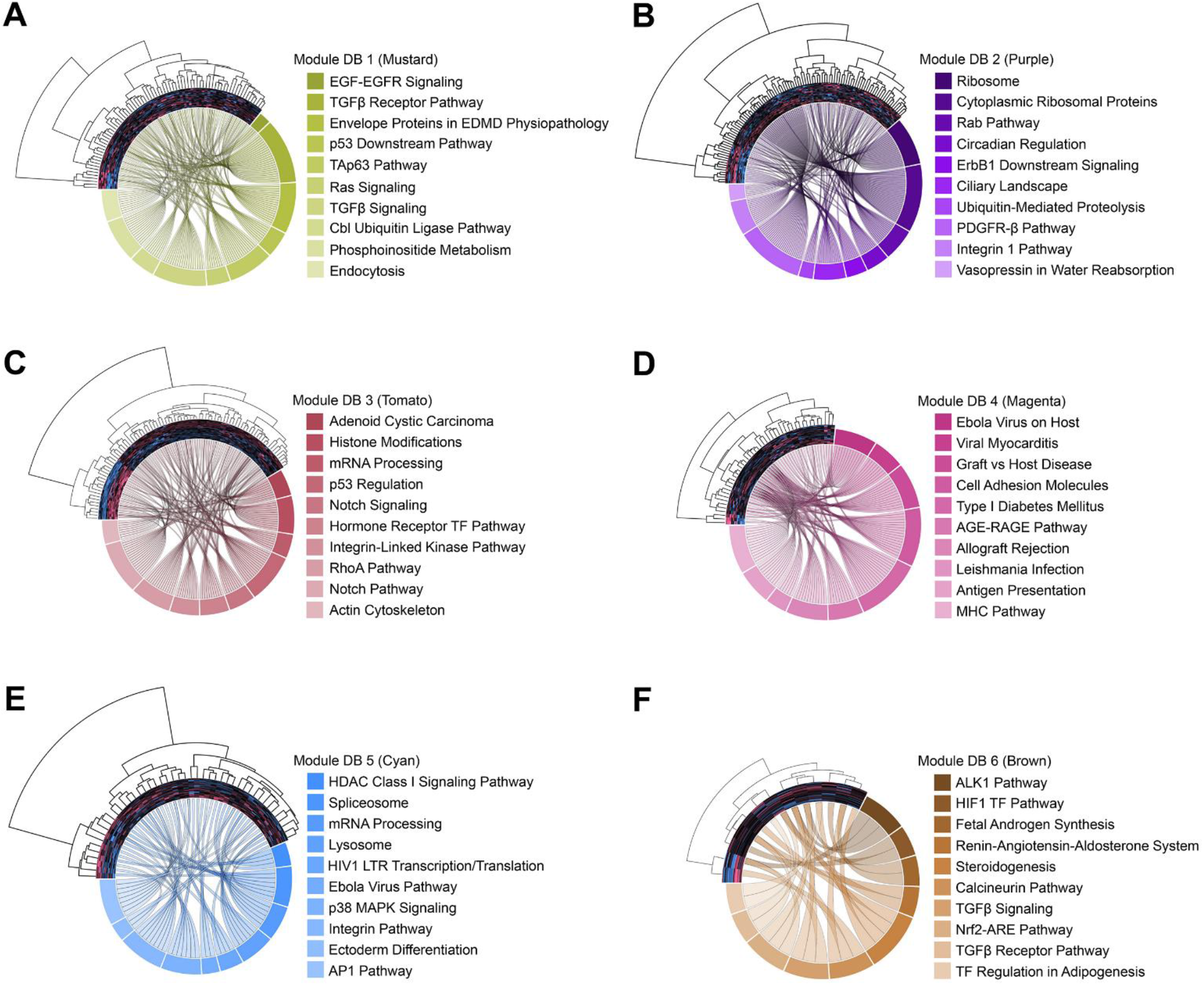
Pathway enrichment for all co-expression modules identified in the decidua basalis (DB) placenta specimens. (A-F) Chord diagrams depicting the top10 enriched pathways associated with the transcripts (depicted as clustered heatmaps) of each of the 6 DB correlation network modules, ranked according to *p*-value. The Gray Module (unassigned transcripts) was not included in this analysis. Each chord represents the connection between an individual module transcript in the heatmap and its presence in a given gene set.

**Figure S6.**
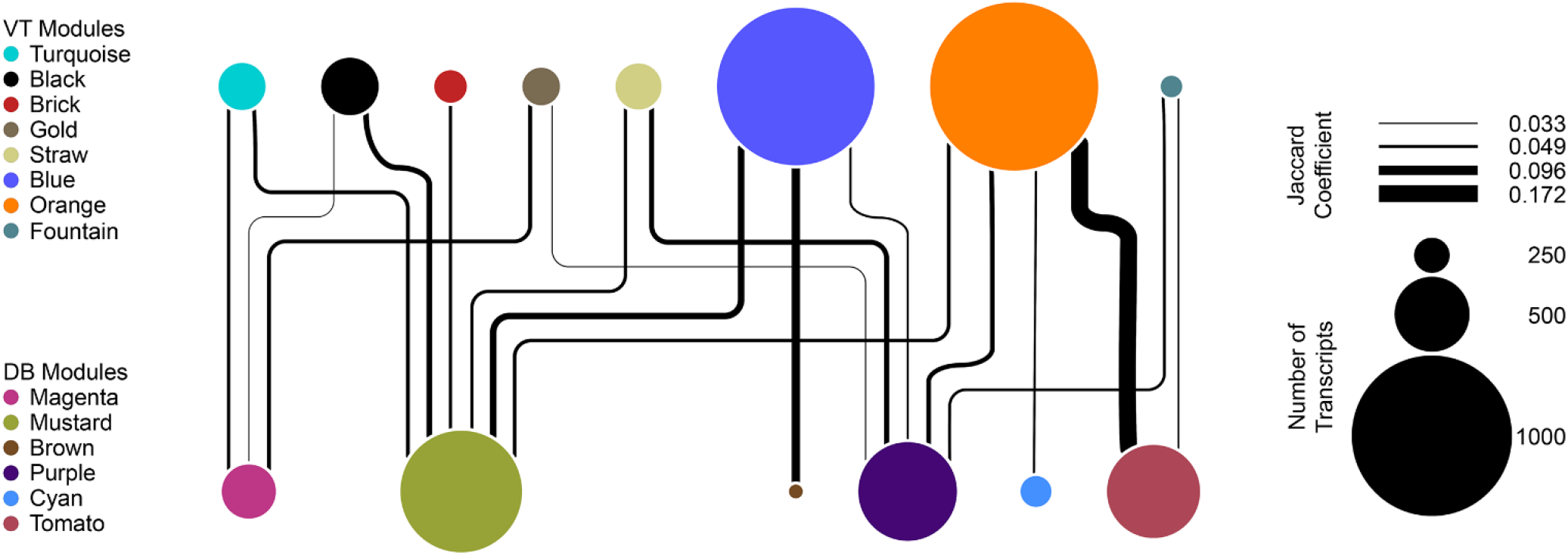
Similarity between co-expression networks of villous tissue (VT) and decidua basalis (DB). Network diagram representing the degree of transcript overlap among co-expression modules identified in each placental region. Node size is proportional to the number of genes within the individual module, identified by color code. Edge connections and their relative sizes represent the degree of transcript overlap between modules, measured using the Jaccard similarity coefficient. Edges were filtered for Jaccard indices <0.032 to clarify the presentation (28 edges omitted). The diagram was constructed using the igraph R package and rendered using Cytoscape v3.9.0.

**Figure S7.**
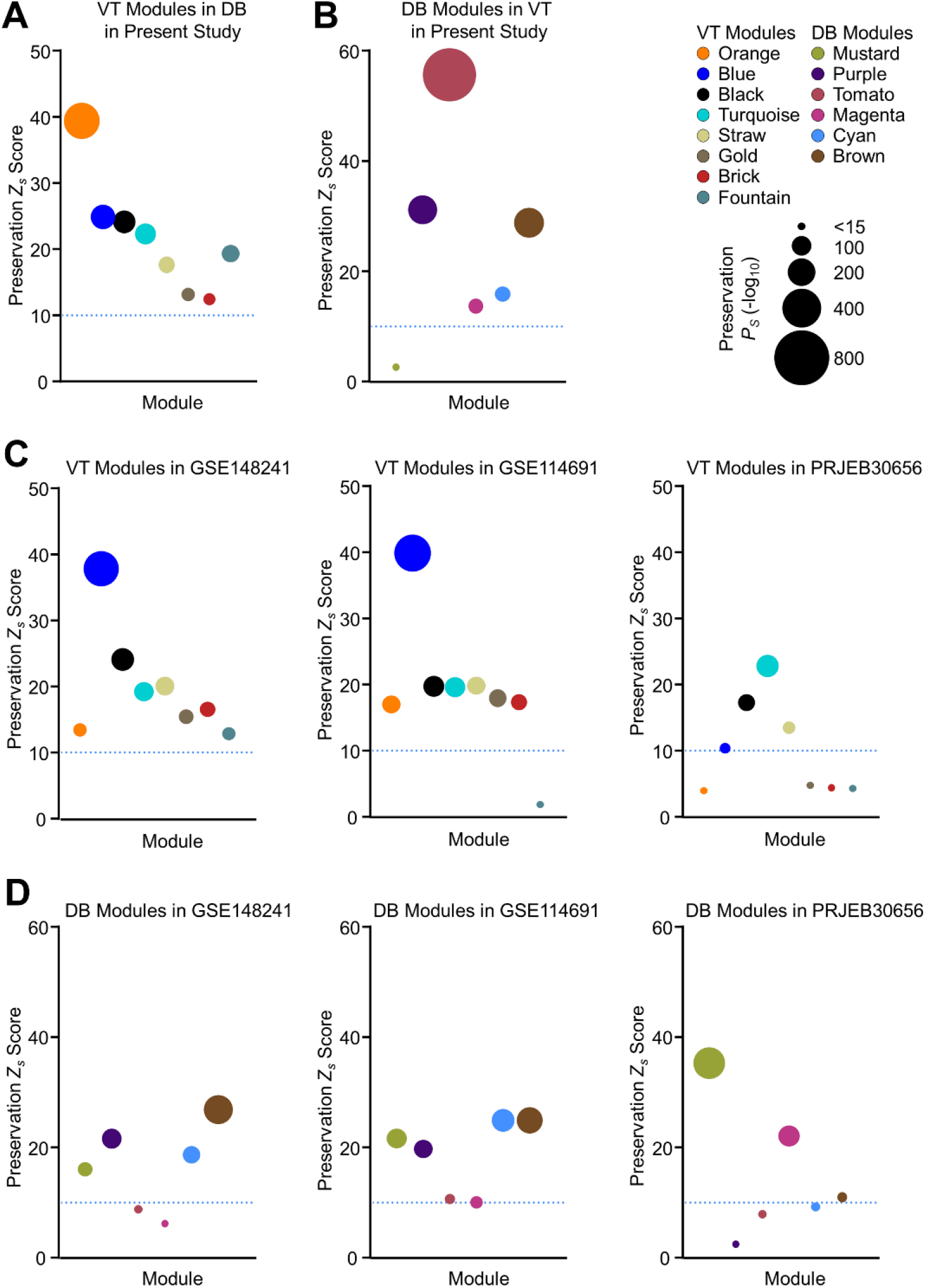
Module preservation between placental regions and in previously published datasets. Bubble plots displaying module preservation summary *Z* statistics (*Z*_s_) for each network module, with data point sizes indicating -log_10_ values for summary *p*-values (*P_s_*). Horizontal blue dotted lines indicate the demarcation between *Z*_s_ scores that indicate strong evidence for module preservation (*Z*_s_>10) and those showing minimal to weak evidence for reproducibility (*Z*_s_<10). (A) Degree of preservation for VT network modules in DB samples in the present study. (B) Preservation of DB network modules in VT specimens in the present study. (C) Preservation of VT network modules in previously published datasets: (left) GSE148241 (n=41); (middle) GSE114691 (n=79); and (right) and PRJEB30656 (n=10). (D) Preservation of DB network modules in GSE148241 (left), GSE114691 (middle), and PRJEB30656 (right) datasets.

**Figure S8.**
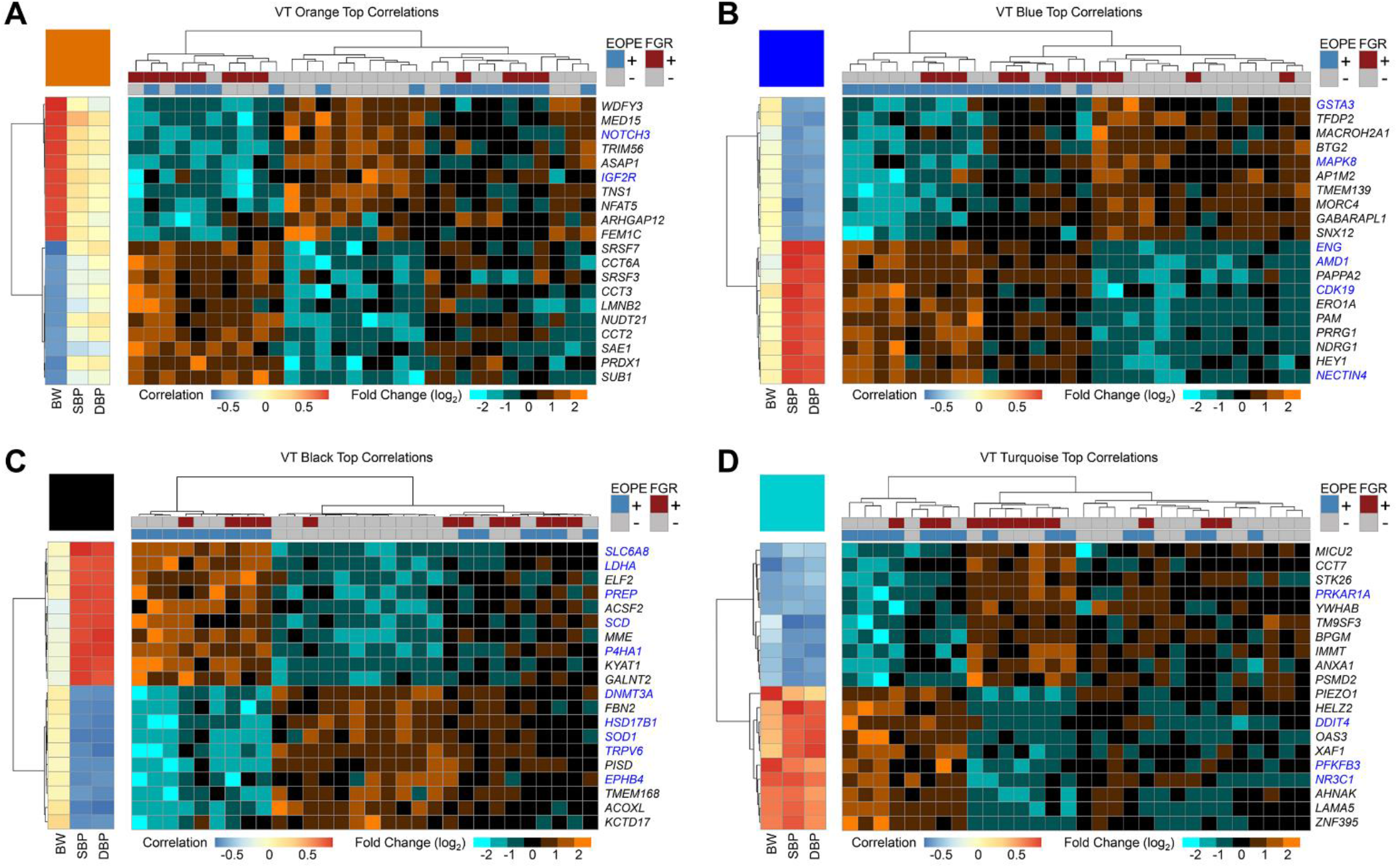
Correlation of individual transcripts with clinical traits for selected co-expression network modules in villous tissue (VT) placental specimens. Heatmaps with unsupervised hierarchical clustering applied to the individual VT specimen transcripts most positively (n=10) and negatively (n=10) correlated with selected clinical variables, as identified through the results of the module-trait correlation analysis overall. Pearson correlation coefficients relating gene expression patterns to BW, SBP, and DBP are shown on the left side of each panel, while row-normalized relative expression data are presented on the right. Color intensity ranges are shown in the keys below the individual heatmaps. Blue text indicates known therapeutic targets documented in the Therapeutic Target Database. (A) Orange VT module, with expressed genes predominantly correlated with BW. (B) Blue VT module, with transcripts chiefly correlated with parental SBP and DBP. (C) Black VT module, primarily correlated with SBP and DBP. (D) Turquoise VT module, with gene expression significantly correlated with BW and DBP. Abbreviations: BW, birthweight; DBP, diastolic blood pressure; EOPE, early-onset severe preeclampsia; FGR, fetal growth restriction; SBP, systolic blood pressure.

**Figure S9.**
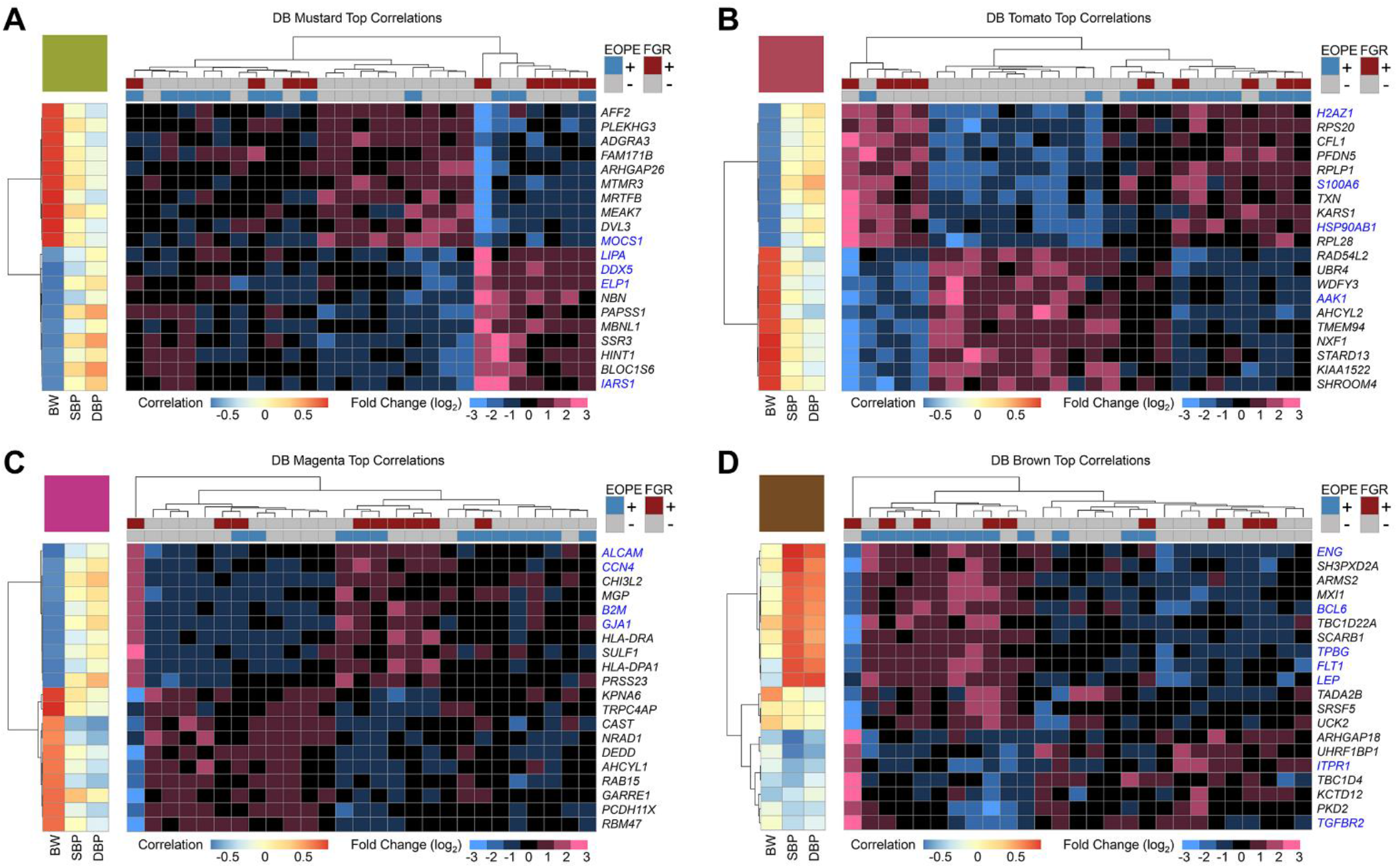
Correlation of individual transcripts with clinical traits for selected co-expression network modules in decidua basalis (DB) placental specimens. Heatmaps with unsupervised hierarchical clustering applied to the DB specimen transcripts most positively (n=10) and negatively (n=10) correlated with selected clinical variables, guided by the results of module-trait correlation analysis overall. Pearson correlation coefficients relating gene expression patterns to BW, SBP, and DBP are shown on the left side of each panel, while row-normalized relative expression data are presented on the right. Color intensity ranges are shown in the keys below individual heatmaps. Blue text indicates known therapeutic targets documented in the Therapeutic Target Database. (A) Mustard DB module, with transcript expression patterns principally correlated BW. (B) Tomato DB Module, mainly correlated with BW. (C) Magenta DB module, with predominant BW correlation. (D) Brown DB module, with significant SBP correlation overall. Abbreviations: BW, birthweight; DBP, diastolic blood pressure; EOPE, early-onset severe preeclampsia; FGR, fetal growth restriction; SBP, systolic blood pressure.

**Figure S10.**
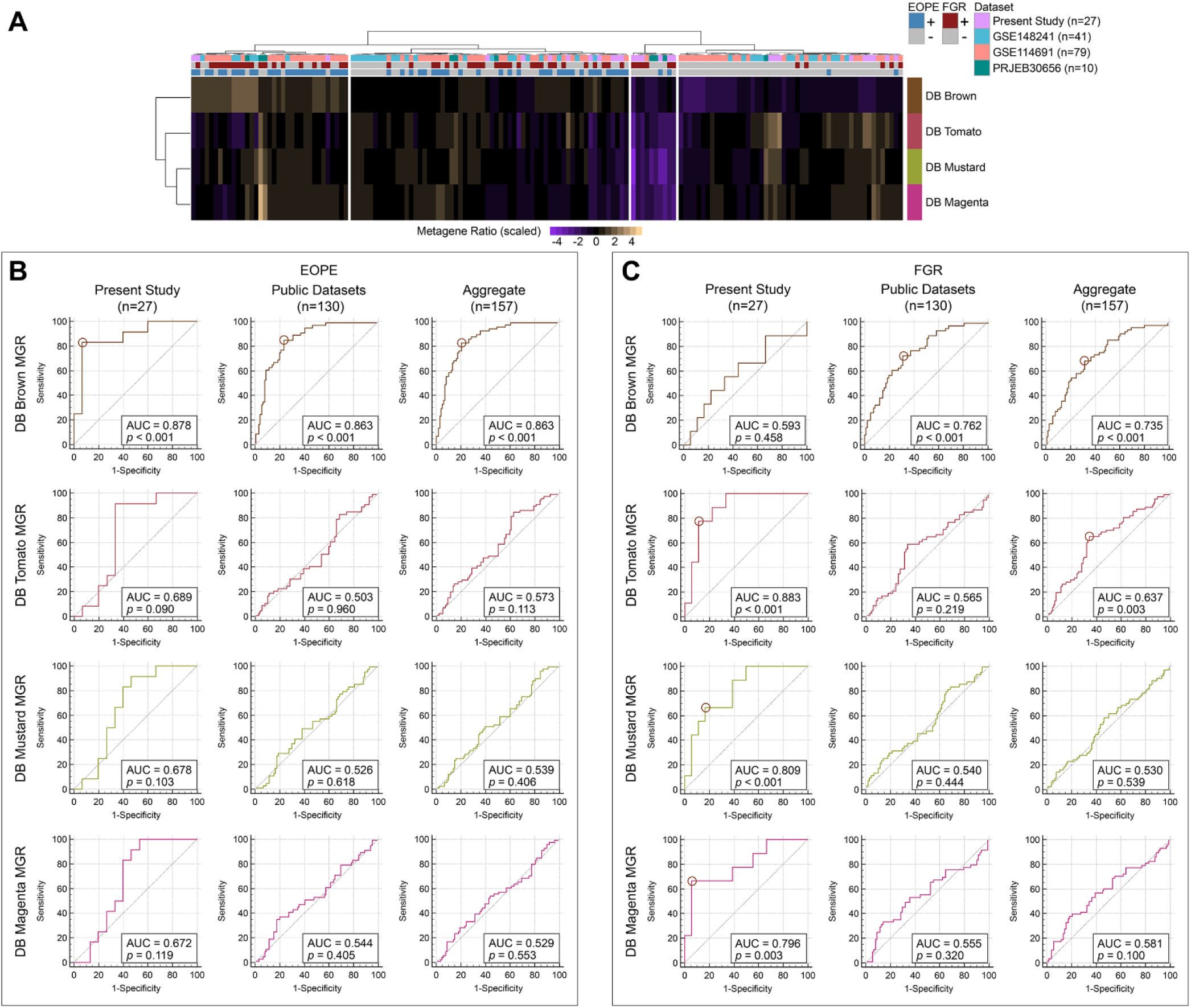
Metagene ratio (MGR) analysis for selected decidua basalis (DB) correlation network modules and associations with clinical diagnoses. (A) Heatmap with unsupervised hierarchical clustering applied to MGRs for the DB co-expression network modules indicated. In this representation, module MGRs have been converted to their corresponding Z scores (based on the mean and standard deviation in each row) to illustrate differences. (B, C) Receiver-operating characteristic (ROC) curves for MGRs of selected DB modules in association with (B) early-onset severe preeclampsia (EOPE) and (C) fetal growth restriction (FGR), based on clinical criteria proximal to the time of delivery. For each ROC curve, sensitivity (true positive rate) is plotted on the y-axis, and 1 - specificity (false positive rate) is plotted on the x-axis. ROCs for the DB specimens in the present study (n=27), public datasets (GSE148241, GSE114691, and PRJEB30656; n=130), and the aggregated set of all samples (n=157) are shown in the leftmost, middle, and rightmost columns of panels B and C, respectively. Each row in panels B and C represents the performance characteristics of the MGR from an individual DB module. The points where Youden’s *J* is maximized (*J*_max_) are indicated by circles in each plot in which the area under the ROC curve (AUC) is significantly different from 0.5 (*p*<0.05).

**Figure S11.**
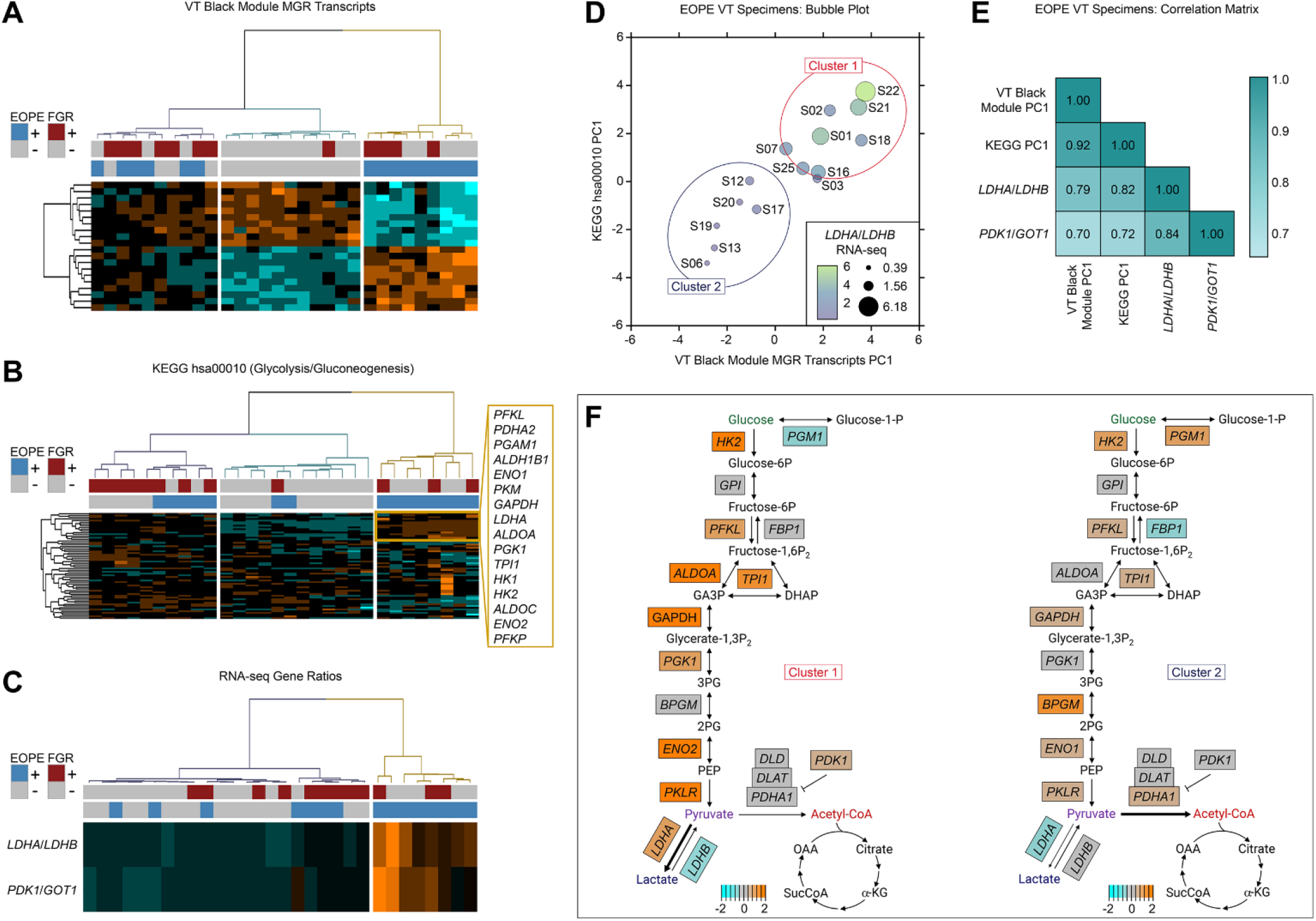
Transcriptional signatures define a molecular subphenotype of EOPE in villous tissue (VT) samples consistent with a glycolytic metabolic shift. (A-C) Heatmaps with unsupervised hierarchical clustering based on: (A) VT Black module transcripts used in the construction of the metagene ratio (MGR) construction; (B) transcripts associated with the KEGG pathway hsa00010 (Glycolysis/Gluconeogenesis); and (C) normalized RNA-seq expression ratios of *LDHA*/*LDHB* and *PDK1*/*GOT1*. (D) Bubble plot comparing the first principal components (PCs) of EOPE±FGR VT specimens stratified by the expression of MGR constituent transcripts from the VT Black module (x-axis) and transcripts mapped to the hsa00010 KEGG pathway (y-axis), both in relation to the expression ratios of *LDHA*/*LDHB* RNA-seq (indicated by bubble size and color). Two molecular clusters could be identified consistently. (E) Matrix displaying Pearson correlation coefficients in pairs comprising the PC1 values for VT Black MGR constituent transcripts, the PC1 values for the KEGG hsa00010 transcripts, and the RNA-seq expression ratios for *LDHA*/*LDHB* and *PDK1*/*GOT1*. All correlations were statistically significant at *p*<0.01. (F) Pathway diagrams adapted from the KEGG hsa00010 map (with the addition of *PDK1* as a critical regulator of the pyruvate dehydrogenase complex) showing the expression of EOPE VT specimen transcripts, subdivided by molecular cluster, relative to iPTB VT samples. Samples from Cluster 1 (left) showed a pattern consistent with a glycolytic metabolic shift toward increased lactate production, while those of Cluster 2 (right) showed expression changes consistent with acetyl-CoA production and entry into the citric acid cycle. Abbreviations: 2PG, 2-phosphoglycerate; 3PG, 3-phosphoglycerate; α-KG, alpha-ketoglutarate; DHAP, dihydroxyacetone phosphate; EOPE, early-onset severe preeclampsia; FGR, fetal growth restriction; GA3P, glyceraldehyde 3-phosphate; OAA, oxaloacetic acid; PC, principal component; PEP, phosphoenolpyruvate; SucCoA, succinyl coenzyme A; VT, villous tissue.

**Figure S12.**
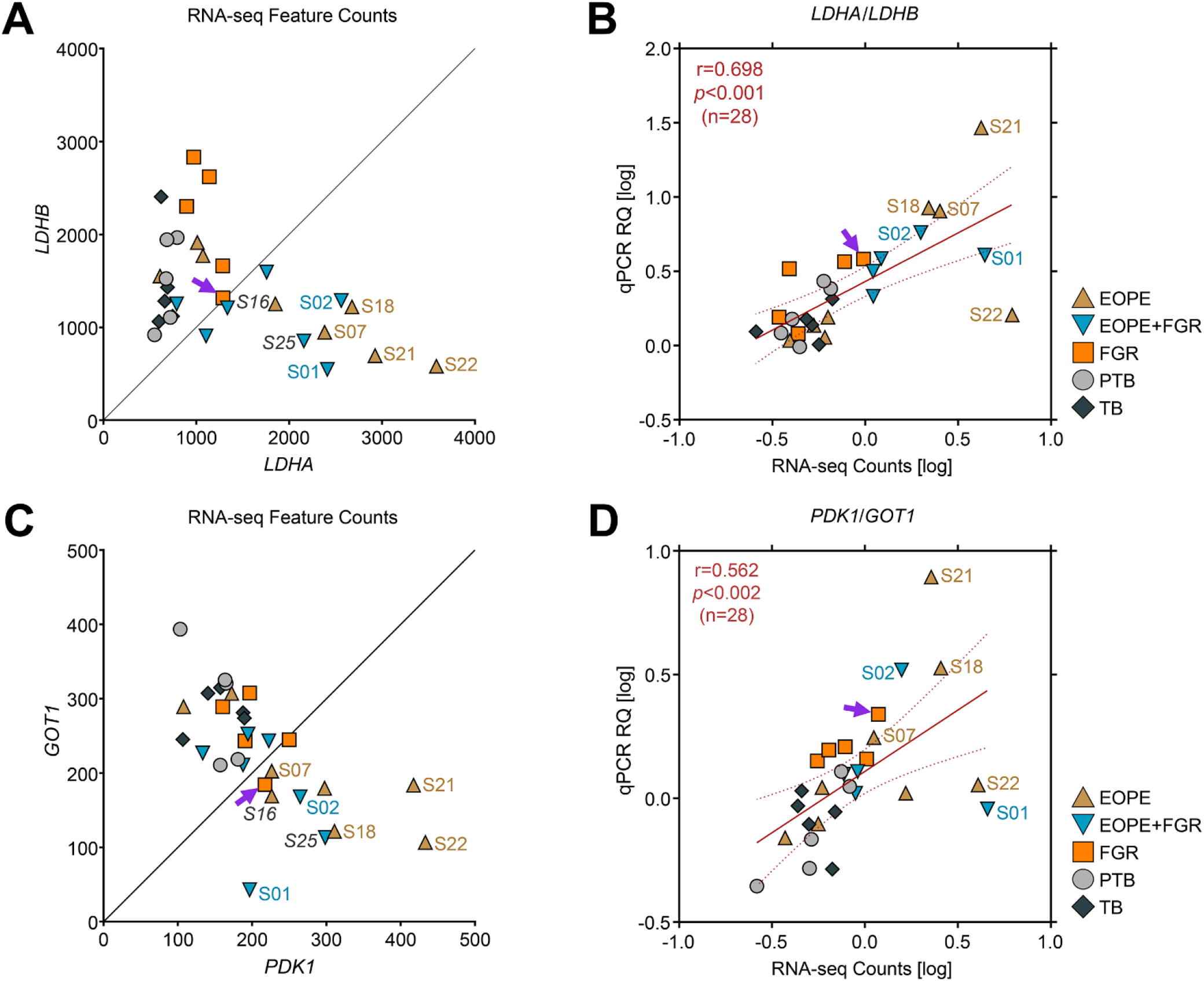
Cross-validation of RNA-seq ratiometric expression by qPCR. (A) Scatterplot showing normalized *LDHA* RNA-seq feature counts in relation to *LDHB* for VT samples (n=30). Samples with molecular evidence for a metabolic shift (Cluster 1 samples in Supplemental Figure S9) are indicated with colored labels. Labels in italics were not included in the qPCR cross-validation. The purple arrow indicates FGR sample S08, which had an elevated *PDK1*/*GOT1* expression ratio. (B) Correlations between the log-transformed *LDHA*/*LDHB* expression ratios from RNA-seq and qPCR experiments were evaluated using Pearson’s product-moment correlation (r=0.698, p<0.001, n=28; samples S16 and S25 were not cross-validated by qPCR due to unavailability of sufficient material). (C) Scatterplot showing normalized RNA-seq feature counts for *PDK1* and *GOT1* in VT samples (n=30). Sample labeling is as in panel A. (D) Correlations between the log-transformed *PDK1*/*GOT1* expression ratios from RNA-seq and qPCR experiments were assessed using Pearson’s product-moment correlation (r=0.562, p<0.002, n=28). Abbreviations: Relative quantification (RQ); VT, villous tissue.

**Figure S13.**
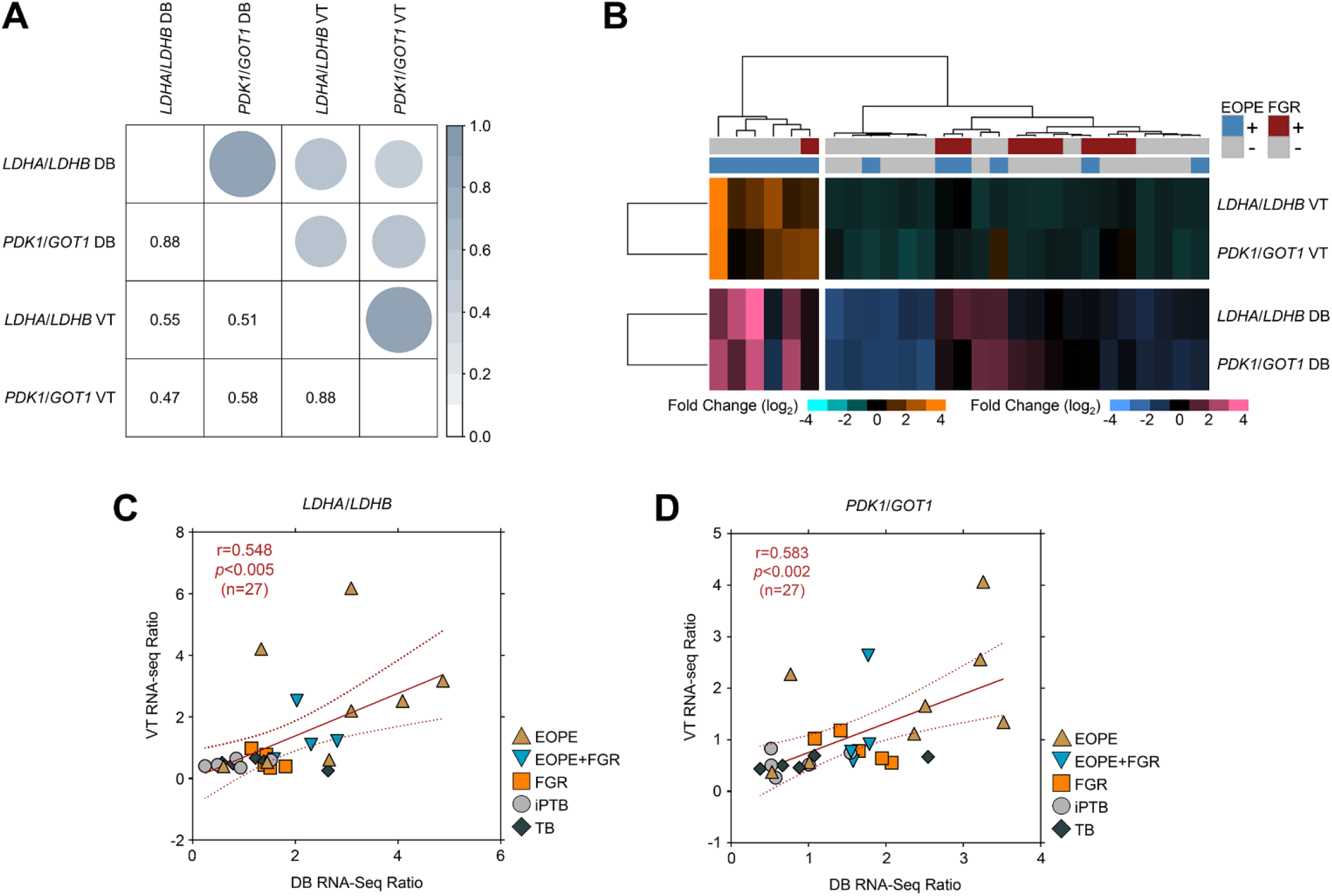
Comparison of the *LDHA*/*LDHB* and *PDK1*/*GOT1* RNA-seq gene expression ratios in paired VT and DB specimens. (A) Correlogram showing the Pearson’s product-moment correlation coefficients among the *LDHA*/*LDHB* and *PDK1*/*GOT1* RNA-seq ratios for the VT and DB anatomical placental regions in paired specimens (n=27). All comparisons were statistically significant (*p*<0.01). The within-region ratio correlations (i.e., DB *LDHA*/*LDHB* vs. DB *PDK1*/*GOT1*; VT *LDHA*/*LDHB* vs. VT *PDK1*/*GOT1*) were stronger than the between-region correlations (i.e., VT *LDHA*/*LDHB* vs. DB *LDHA*/*LDHB*; VT *PDK1*/*GOT1* vs. DB *PDK1*/*GOT1*). Note that the correlation coefficient for the VT *LDHA*/*LDHB* vs. VT *PDK1*/*GOT1* comparison differs from that in Figure S11E since the latter contains additional VT samples, S01, S02, and S03, for which paired DB samples were unavailable. (B) Heatmap showing the distribution of samples following unsupervised hierarchical clustering based on the combined dataset comprising the *LDHA*/*LDHB* (panel C) and *PDK1*/*GOT1* normalized RNA-seq ratios for the VT and DB placental anatomical regions. (C, D) Scatterplots showing the sample distribution obtained when comparing the normalized *LDHA*/*LDHB* (panel C) and *PDK1*/*GOT1* (panel D) RNA-seq ratios between the VT and DB placental regions. Abbreviations: DB, basal plate decidual basalis; VT, villous tissue.

**Figure S14.**
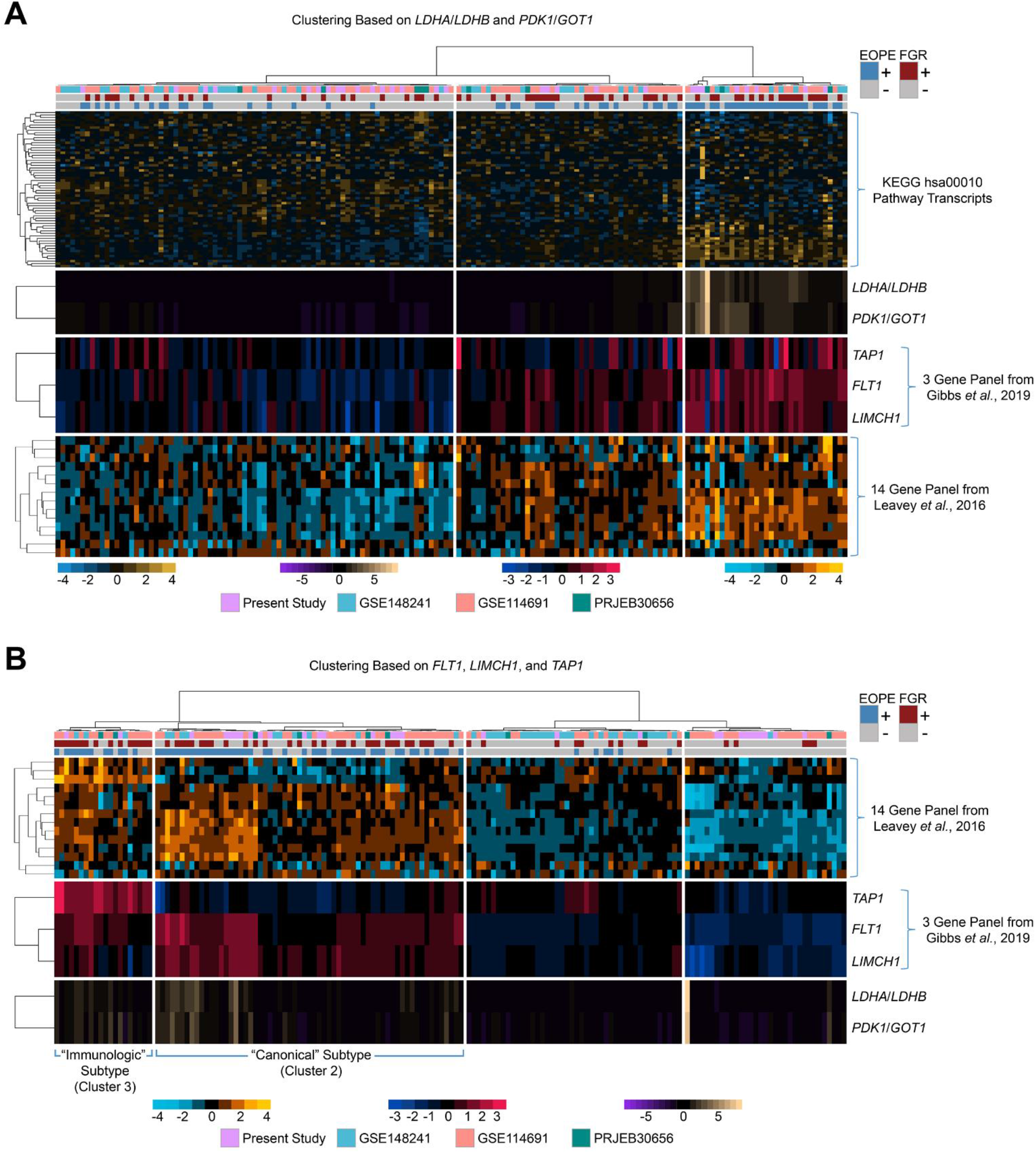
Relationships between the transcriptional signatures suggestive of a glycolytic metabolic shift and those associated with molecular subtypes of PE and FGR identified in prior studies. Heatmaps, aligned by columns, showing the transcriptional expression patterns in individual VT samples from each of the four studies (the present study and the 3 previously published datasets, n=160). The 14 gene panel was abstracted from the supplementary materials in Leavey *et al.* (Hypertension 2016;68:137-47) and consisted of candidate transcripts used in the initial evaluation of a qPCR-based molecular classification strategy for PE (*ENG*, *FLT1*, *FSTL3*, *LIMCH1*, *MAN1C1*, *METTL18*, *MORN3*, *MT1F*, *PIK3CB*, *SNX10*, *SQOR*, *TAP1*, *TPBG*, and *VPS54*). The 3 genes used for the qPCR-based classification algorithm in Gibbs *et al*. (Am J Obstet Gynecol 2019;220:110.e1- 110.e21) comprised a subset of this larger panel. The same data have been arranged by unsupervised hierarchical clustering based on: (A) the expression ratios of *LDHA*/*LDHB* and *PDK1*/*GOT1*; and (B) the 3 gene panel from the Gibbs *et al*. publication (*FLT1*, *LIMCH1*, and *TAP1*). These arrangements are intended to illustrate the relationships between the molecular subtypes of PE and FGR identified previously (emphasized in panel B) and the samples exhibiting evidence of a metabolic shift in the present study (emphasized by the arrangement in panel A). Expression patterns that approximately correlate with clusters 2 and 3 as described in Leavey *et al.* (Hypertension 2016;68:137-47) and Gibbs *et al*. (Am J Obstet Gynecol 2019;220:110.e1-110.e21) are indicated in panel B. Abbreviations: EOPE, early-onset severe preeclampsia; FGR, fetal growth restriction; VT, villous tissue.

## Major Resources Table

In order to allow validation and replication of experiments, all essential research materials listed in the Methods should be included in the Major Resources Table below. Authors are encouraged to use public repositories for protocols, data, code, and other materials and provide persistent identifiers and/or links to repositories when available. Authors may add or delete rows as needed.

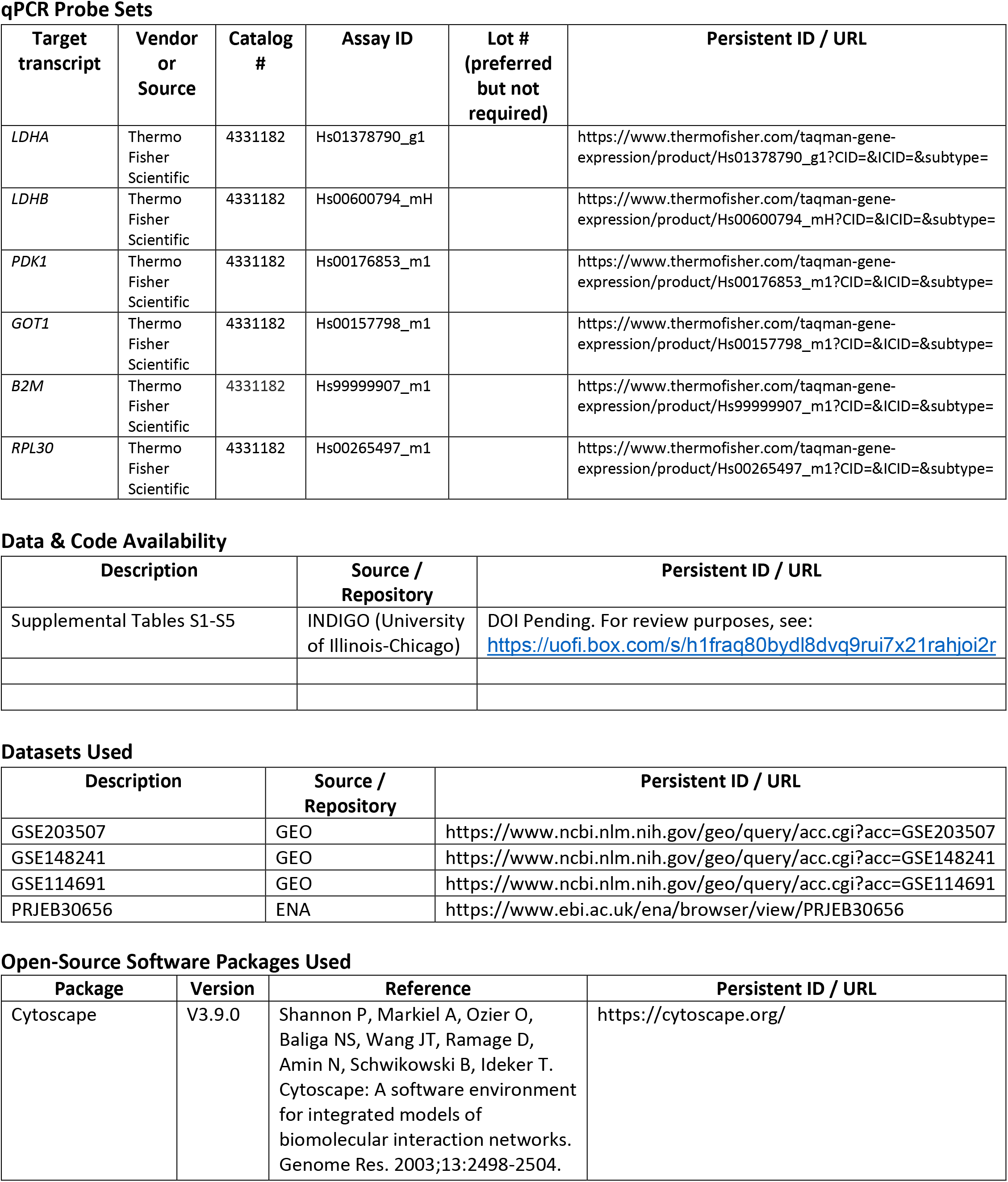
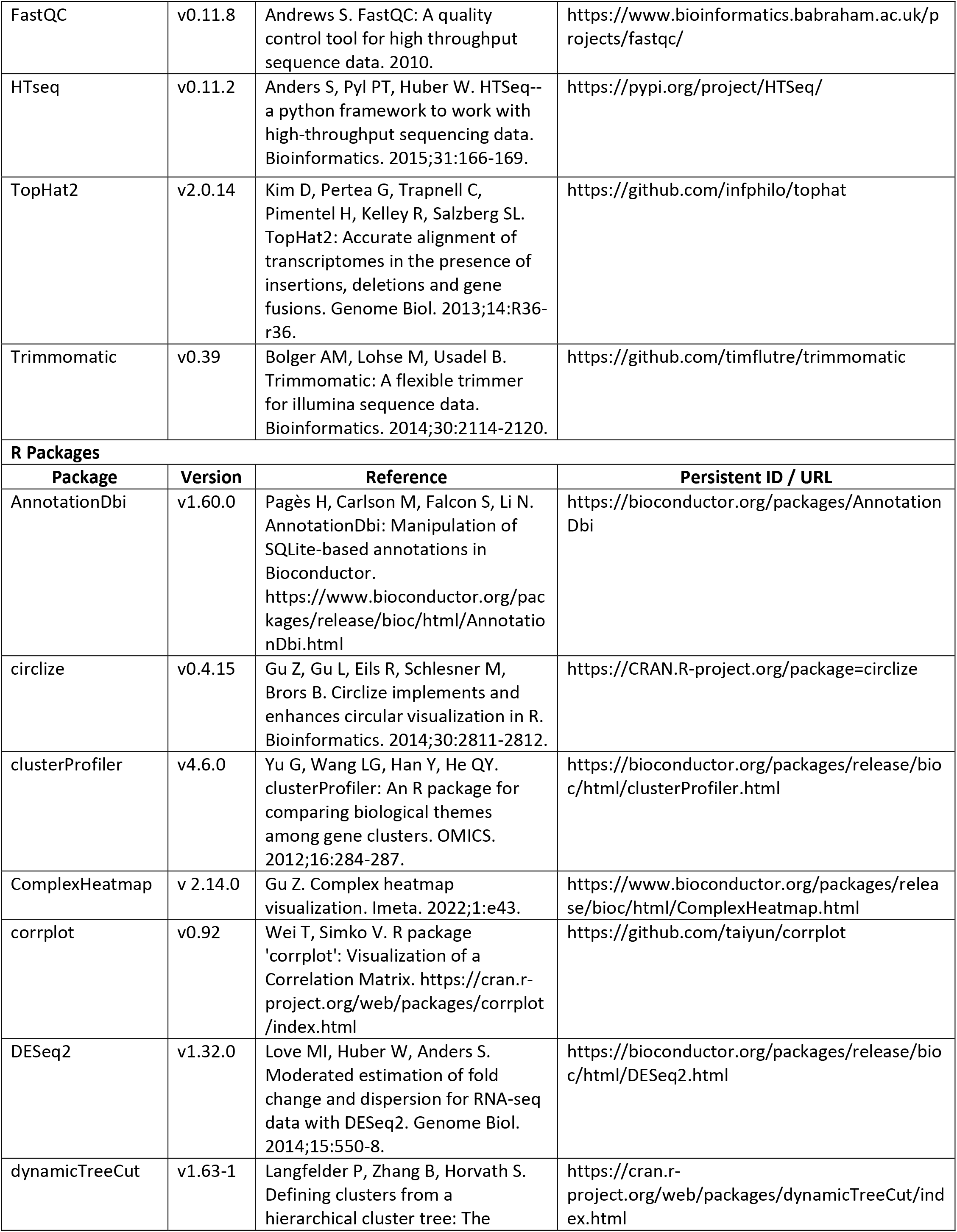
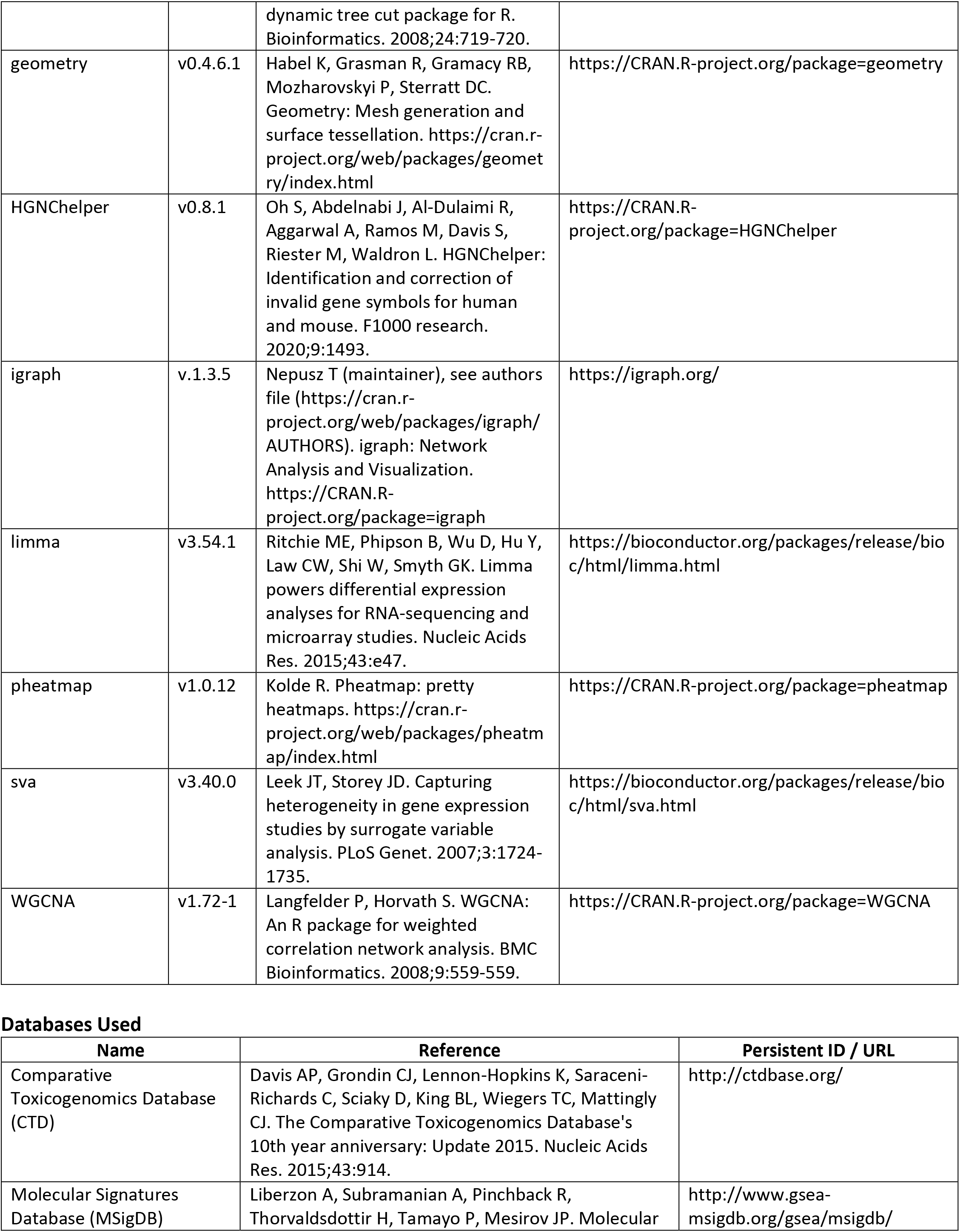
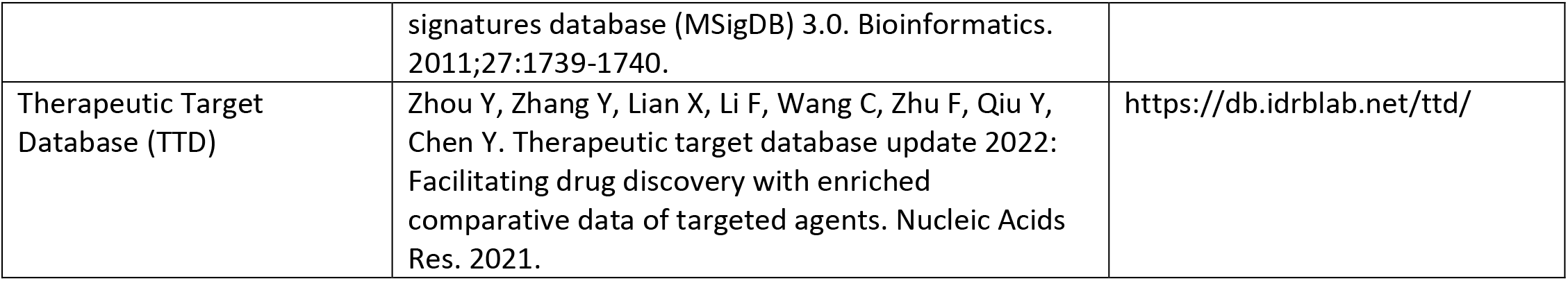

